# Multimodal evidence for a mechanistic model of working memory deficits in schizophrenia

**DOI:** 10.64898/2026.06.24.26356367

**Authors:** Tuomo Mäki-Marttunen, Nadine Parker, Verónica Mäki-Marttunen, Samuel Neymotin, Alexey Shadrin, Ibrahim Akkouh, Linn Sofie Sæther, Torill Ueland, Marja-Leena Linne, Torbjørn Elvsåshagen, Srdjan Djurovic, Ole A. Andreassen, Gaute T. Einevoll

**Affiliations:** Faculty of Medicine and Health Technology, Tampere University, Tampere, Finland; Department of Biosciences, University of Oslo, Oslo, Norway; Centre for Precision Psychiatry, Division of Mental Health and Addiction, Oslo University Hospital & Institute of Clinical Medicine, University of Oslo, Oslo, Norway; Center for Biomedical Imaging and Neuromodulation, Nathan S. Kline Institute for Psychiatric Research, Orangeburg, NY, USA; Department Psychiatry, NYU Grossman School of Medicine, New York, NY, USA; Department of Medical Genetics, Oslo University Hospital & University of Oslo, Oslo, Norway; Section for Clinical Psychosis Research, Division of Mental Health and Addiction, Oslo University Hospital, Oslo, Norway; Department of Psychology, University of Oslo, Oslo, Norway; Department of Neurology, Division of Clinical Neuroscience, Oslo University Hospital, Oslo, Norway; Department of Behavioural Medicine, Institute of Basic Medical Sciences, Faculty of Medicine, University of Oslo, Oslo, Norway; Department of Physics, Norwegian University of Life Sciences, Ås, Norway; Department of Physics, University of Oslo, Oslo, Norway

## Abstract

Working memory (WM) deficits are central to schizophrenia (SCZ), yet their mechanistic basis remains unclear. We combined computational modelling with genetic, transcriptomic, behavioural, and fMRI data to construct a mechanistic account of WM impairment in SCZ. Post-mortem RNA expression from prefrontal and anterior cingulate cortex (ACC) was integrated with single-cell, network, and synaptic plasticity models to show how SCZ-related changes in ion channel-encoding and plasticity-regulating genes alter sustained delay-period activity and long-term potentiation, suggesting an impairment of WM. The model predictions were supported by behavioural WM test (letter-number sequencing) results and polygenic risk scores for SCZ based on ion channel and plasticity gene sets. Mendelian randomization, together with nominally significant single-gene risk analyses, implicated specific ion channel genes, particularly CACNA1I, as putatively causal for both SCZ liability and WM deficits. fMRI N-back data supported ACC-specific delay-period impairments. These multimodal findings highlight candidate, druggable mechanisms for cognition-focused interventions in SCZ.

## 1 Introduction

Working memory (WM) deficits are a core feature of schizophrenia (SCZ) and contribute substantially to functional and occupational impairment, yet their mechanisms remain poorly understood. Patients with SCZ show impairments across WM tasks and paradigms, beyond generalized cognitive dysfunction [Lee and Park, 2005, Forbes et al., 2009, Barch and Ceaser, 2012]. Current antipsychotic medications have limited effects on cognitive symptoms, including WM, which remain a major unmet therapeutic need [Keefe and Harvey, 2012, Green, 2016]. At the same time, large-scale genetic studies firmly establish SCZ as a highly polygenic disorder enriched for synaptic and neuronal genes [Ripke et al., 2014, Devor et al., 2017, Trubetskoy et al., 2022], and post-mortem transcriptomic analyses report coordinated changes in expression of ion channel-encoding and plasticity-regulating genes in prefrontal and anterior cingulate cortex (PFC, ACC) of patients [Hoffman et al., 2019, Mäki-Marttunen et al., 2024]. However, it remains unclear how such polygenic and regionally specific expression changes give rise to disturbances in neuronal excitability, synaptic plasticity, and sustained delay-period activity that support WM [Miller et al., 1996].

In the present study, we applied computational modelling, polygenic and single-gene risk scores, and Mendelian randomization (MR) and four data modalities (post-mortem expression data, behavioural WM test results, GWAS risk data, and blood-oxygen-level-dependent (BOLD) signal data) to propose a mechanism through which WM deficits in SCZ partly occur (Fig. 1). First, we integrated post-mortem cortical RNA expression data from patients and controls with 1) single-cell and network models of pyramidal cells and interneurons [Neymotin et al., 2016, Mäki-Marttunen et al., 2024] to identify ion channel-mediated mechanisms for WM impairment and 2) a biochemically detailed single-synapse model of synaptic plasticity to test if the synaptic plasticity deficits observed in [Mäki-Marttunen et al., 2024] correlate with these mechanisms. We then related the resulting alterations in sustained firing to WM performance and polygenic risk scores (PRSs) of SCZ based on ion channel-encoding and plasticity-regulating genes using genetic and behavioural (letter-number sequencing; LNS) data from the Thematically Organized Psychosis (TOP) study [Haatveit et al., 2023]. Finally, using single-gene SCZ PRS and bidirectional MR analyses on brain expression quantitative trait locus (eQTL) data [De Klein et al., 2023, Hemani et al., 2018], we tested whether genetically proxied expression of specific ion-channel genes shows putative causal relationships with SCZ liability and whether the SCZ risk within these genes is associated with WM performance. Our analysis provides a plausible mechanism by which altered expression of ion channel-encoding genes in the ACC can weaken the delay-period activity in cortical circuits and impair WM — the expression data from the ACC compared with modelling of postsynaptic plasticity pathways also suggests that these effects are associated with an increase in basal synaptic strength and a compromised long-term potentiation (LTP) amplitude. The predictions of ACC-specific impairments of delay-period activity were supported by results from an open functional magnetic resonance imaging (fMRI) dataset [Repov̌s and Barch, 2012] recorded during an N-back WM task from healthy controls (HCs) and participants with SCZ. Our multimodal results converge to suggest that CACNA1I single nucleotide polymorphisms (SNPs), and the increased CACNA1I expression that they are likely to cause, contribute to both SCZ risk and WM impairment. These findings can lead to identification of druggable targets to treat cognitive symptoms in genetics-based subgroups of SCZ.

**Figure 1:**
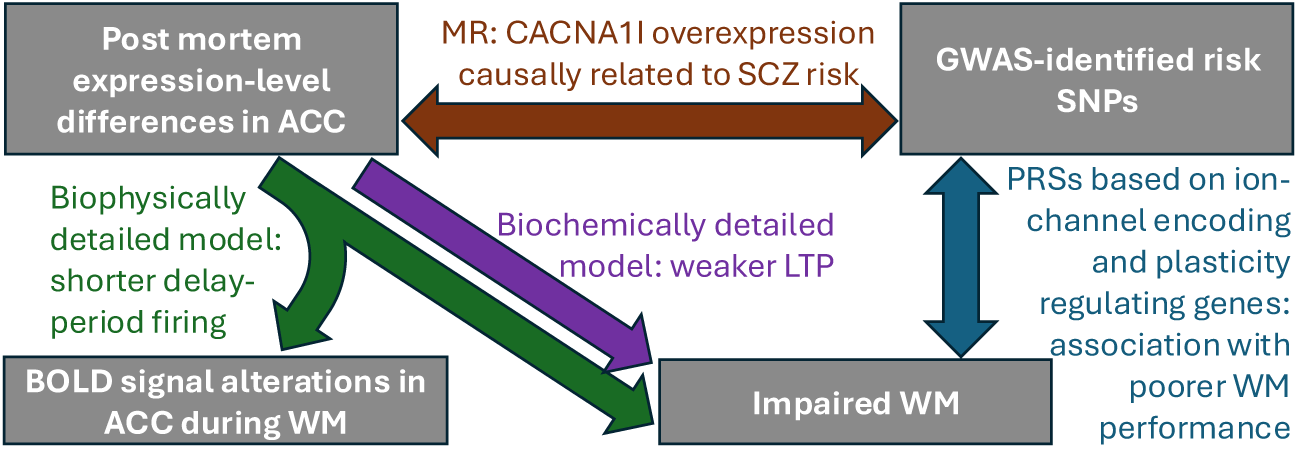
Our study combines four types of data: post-mortem expression data, behavioural WM test results, SNP-wise GWAS risk data, and BOLD imaging. We present multimodal evidence for a mechanism for WM deficits in SCZ where altered expression of genes encoding ion channel subunits or synaptic plasticity-associated genes in ACC leads to a compromised network response to a WM-inducing stimulus and weakened LTP.

## 2 Results

### 2.1 Altered expression of ion channel-encoding SCZ-associated genes predicts a decreased amplitude of the single-neuron response to the WM-inducing stimulus in ACC

We simulated the activity of single pyramidal cells before, during and after a WM-inducing stimulus (a 100-ms stimulation of glutamatergic receptors leading to a long-lasting increase in pyramidal cell excitability) in the HC case and under ion-channel manipulations corresponding to the gene-expression alterations observed in the PFC and ACC in SCZ patients. We used the model of [Neymotin et al., 2016] with a few adjustments (see Methods and Fig. 8) for this and implemented the changes suggested by the psychiatric post-mortem gene expression data set [Hoffman et al., 2019] (CommonMind; see Table 1). Our model predicted that in the PFC the underexpression of SCN1B (affecting the fast Na^+^ currents) causes the response to the WM-inducing stimulus to weaken (Fig. 2A) and that the overexpression of HCN1 (affecting the *I_h_* current) has an opposite, strengthening effect on the response (Fig. 2B). The overexpression of CACNA1I (affecting the T-type Ca^2+^ current) in the PFC had little effect on the single-neuron activity (Fig. 2D), while that of KCNB1 (affecting the delayed-rectifier current) mildly weakened the predicted response (Fig. 2E). In the ACC, the overexpression of KCND3 (affecting the A-type current; Fig. 2C) and, to a lesser extent, that of CACNA1I and KCNB1 (Fig. 2D–E) weakened the response to the WM-inducing stimulus. In both ACC and PFC, the altered expression of ATP2A2, KCNMA1, KCNQ3, CACNA1C, CACNA1D, and KCNJ6 had little or no effect on the response to the WM-inducing stimulus (Fig. S1). When all these variants were combined, the predicted response to the WM-inducing stimulus was decreased for the ACC-like but relatively unchanged for the PFC-like alterations of expression seen in SCZ (Fig. 2F). We confirmed the significance of these results using subject-wise simulations, where the altered model parameters (ion-channel conductances of the fast Na^+^, HCN, T-and L-type Ca^2+^, and A-type, delayed-rectifier, BK, M-type, and GABA_B_R-mediated K^+^ channels, and the SERCA pump efficacy) were determined for each subject based on their RNA expression normalized by the average RNA expression of the corresponding genes among the controls. Each personalized model (N=426 for PFC, N=478 for ACC) was simulated and the time the firing activity remained above twice the HC baseline (the average firing rate during the last three seconds before the WM-inducing stimulus) was determined for each subject. The duration of sustained firing was significantly shorter in the SCZ group than in the HCs in the ACC (p=3.7*×*10*^−^*^7^ *<*0.05, U-test; Fig. 2G) but unchanged in the PFC (p=0.86*>*0.05, U-test; Fig. 2H).

**Figure 2:**
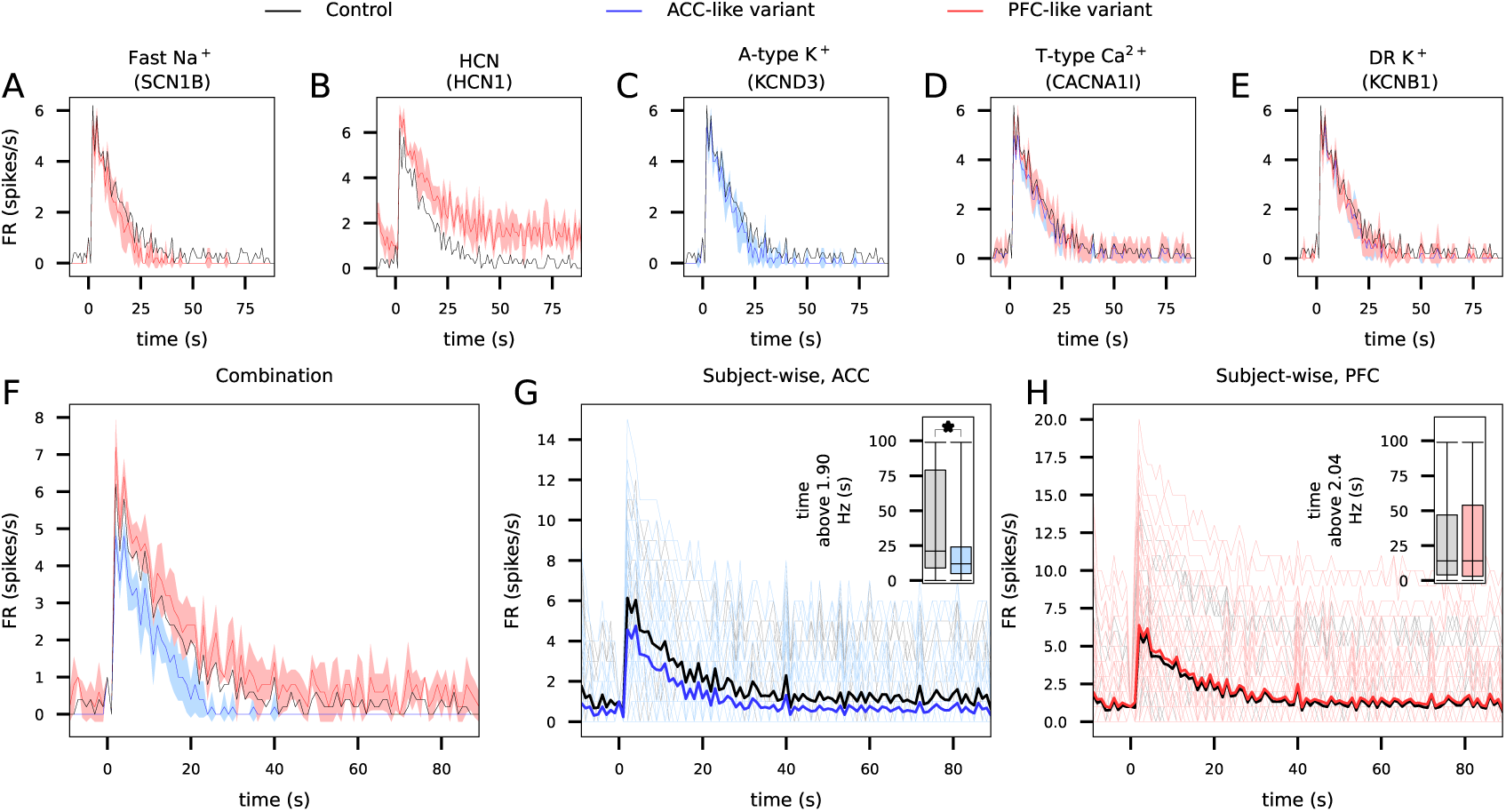
ACC-like variants of ion-channel expression decrease the pyramidal cell response to the WM-inducing stimulus. **A–E**: The simulation of the response to a strong AMPAR, NMDAR and mGluR stimulation (see Fig. 8F and Methods) was repeated under alterations of a single SCZ-associated ion channel as suggested by post-mortem expression data from PFC (red) and/or ACC (blue). Namely the conductance of the fast Na^+^ channel (A), HCN channel (B), A-type K^+^ channel (C), T-type Ca^2+^ channel (D), or delayed-rectifier K^+^ channel (E) was altered according to the expression data of the underlying SCZ-associated genes. **F**: The experiment was repeated by applying all above PFC-(red) or ACC-specific (blue) alterations. **G–H**: The subject-wise simulation results according to expression data from the ACC (G) or PFC (H). The dim gray (HC) and blue (SCZ, ACC) or red (SCZ, PFC) curves show the results from simulations of *personalized models* where the model parameters were tuned based on the subject’s gene expression data relative to an average control. The thick black curves show the mean simulation results across the HC subjects (N=251 in ACC, N=215 in PFC), and the thick blue (ACC, N=227) and red (PFC, N=211) curves show those across the SCZ subjects. Insets: Box plots of the time that the FR was above two times the mean baseline FR in HC subjects on average (1.9 spikes/s in ACC, 2.04 spikes/s in PFC) following a WM-inducing response. The asterisk denotes significantly different time between HC and SCZ subjects (U-test, p*<*0.05).

**Table 1:**
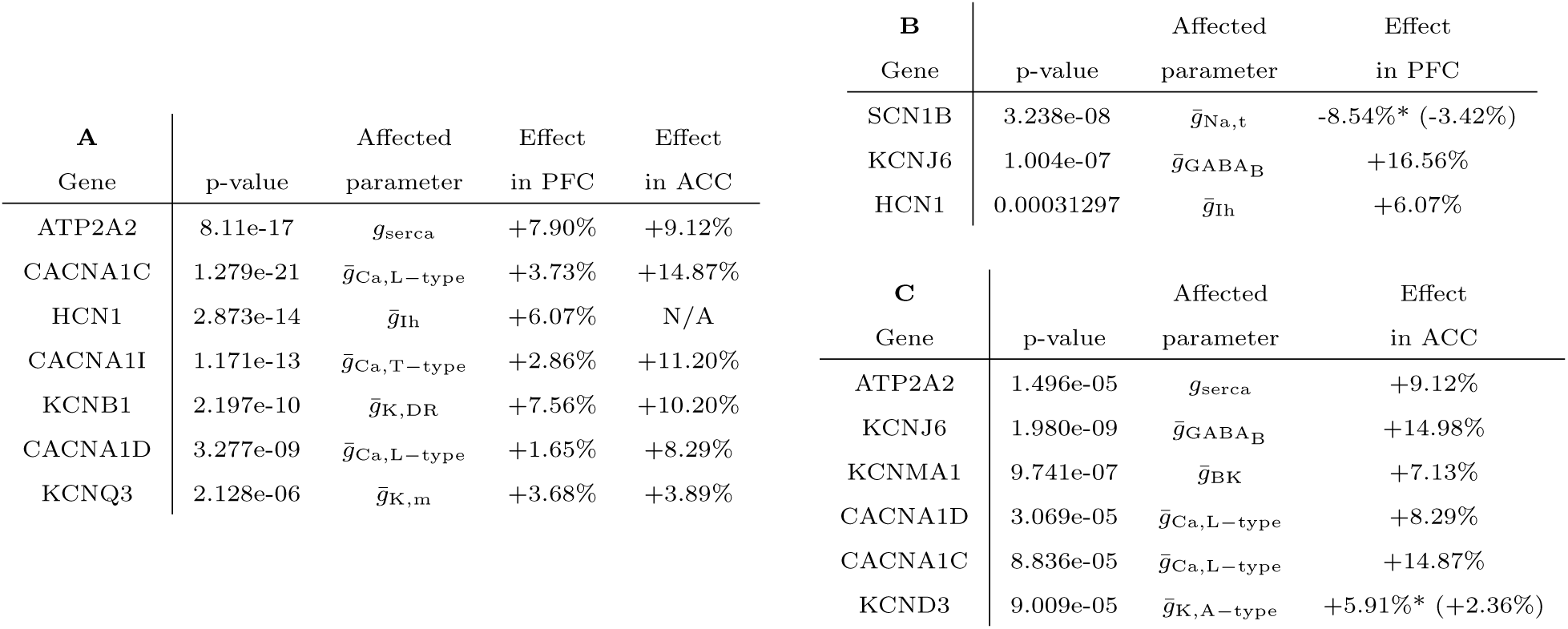
Table of ion channel-encoding genes associated with SCZ and the alterations in expression level in SCZ compared to controls according to post-mortem expression data in PFC and ACC. **A**: Genes that included SNPs moderately or highly associated (p-value *<* 5*×*10*^−^*^6^) with the risk of SCZ according to GWAS data [Trubetskoy et al., 2022]. The second column indicates the minimal p-value among the SNPs of the gene in the data of [Trubetskoy et al., 2022], and the third column shows the model parameter affected by the gene. The fourth and the fifth columns show the average effect of the SCZ diagnosis on the post-mortem expression of the gene in PFC and ACC in the CommonMind data [Hoffman et al., 2019], fitted using a linear model that corrected for the sex, age, and the post-mortem index. **B–C**: Genes whose post-mortem expression in the PFC (B) or ACC (C) was moderately or highly associated (p-value *<* 5*×*10*^−^*^5^) with SCZ according to the CommonMind data [Hoffman et al., 2019]. The p-values in the second column indicate the effect of SCZ on the expression of the gene, and the third and fourth columns show the affected model parameter and the average effect of the SCZ diagnosis on the expression as in (A). (*): The knock-out of genes SCN1B and KCND3 showed only a 40% decrease in the underlying current amplitudes in pyramidal cells [Hull et al., 2020, Norris and Nerbonne, 2010]. We thus apply effects on the conductance parameters that are down-scaled in proportion (in parentheses) in this work.

Taken together, pyramidal neuron spiking responses to a WM-inducing stimulus were decreased in models that were tuned to post-mortem expression data from ACC in SCZ patients but not in those tuned to data from PFC.

### 2.2 Sustained circuit activity after WM-inducing stimulus in ACC is decreased in amplitude in SCZ

The altered activity of single neurons suggests that the sustained network activity following a WM-inducing stimulus may be altered by changes in ion channel expression as well. Here, we simulated the activity of the multi-layer cortical network consisting of 776 neurons in response to a WM-inducing stimulus in the presence and absence of SCZ-associated gene-expression alterations in the pyramidal cells. We measured the average spiking activity over time in the four populations: stimulated (291) and non-stimulated (291) pyramidal cells, and parvalbumin-positive (97) and low-threshold-spiking (LTS) interneurons (97).

Similarly to the firing activity of single pyramidal neurons after WM-inducing stimulus, the network firing activity in response to the WM-inducing stimulus was decreased in the stimulated pyramidal cell population in the SCZ population when tuned with the expression data from the ACC but not the PFC (Fig. 3A). In particular, the duration of the sustained firing in the pyramidal cells (firing rate *>* 2 times the average baseline firing rate in stimulated pyramidal neurons of HCs) was significantly shorter in the ACC-like variants (p=1.1 *×* 10*^−^*^7^ *<*0.05, U-test; Fig. 3B), but unchanged in the PFC-like variants (p=0.89*>*0.05, U-test; Fig. 3C) in the personalized models. Similarly, the duration of sustained firing in the interneurons — both parvalbumin-positive and LTS neurons — was significantly shorter for the ACC but not the PFC variants (ACC, parvalbumin-positive: p=2.6*×*10*^−^*^7^ *<*0.05, U-test, Fig. 3E; PFC, parvalbumin-positive: p=0.87*>*0.05, U-test, Fig. 3F; ACC, LTS: p=1.1*×*10*^−^*^6^ *<*0.05, U-test, Fig. 3H; PFC, LTS: p=0.54*>*0.05, U-test, Fig. 3I). Gene-wise variant simulations show that, similar to single-neuron activity, the predicted SCZ-associated alterations in network activity were mostly driven by underexpression of SCN1B (which weakened the response to the WM-inducing stimulus) and overexpression of HCN1 (which strengthened the response) in the PFC, by overexpression of KCND3 and CACNA1I in the ACC (both of which weakened the response; Fig. S2B–E), and by overexpression of KCNB1 in both regions (which weakened the response; Fig. S2F). Other alterations had little effect (Fig. S2G–K). The suppressing effects of CACNA1I overexpression were mediated by increased Ca^2+^ concentrations in the apical dendrite (but not soma; Fig. S2M–N), which led to larger small-conductance Ca^2+^-activated K^+^ (SK) currents, consistent with our previous model predictions [Mäki-Marttunen et al., 2017, Mäki-Marttunen et al., 2019]. We confirmed this by running the simulations of SCZ-associated alterations of Ca^2+^ channel expression in the total absence of SK currents (Fig. S2O–R) — in these simulations, the overexpression of CACNA1I did not decrease the response to a WM-inducing stimulus (Fig. S2P).

**Figure 3:**
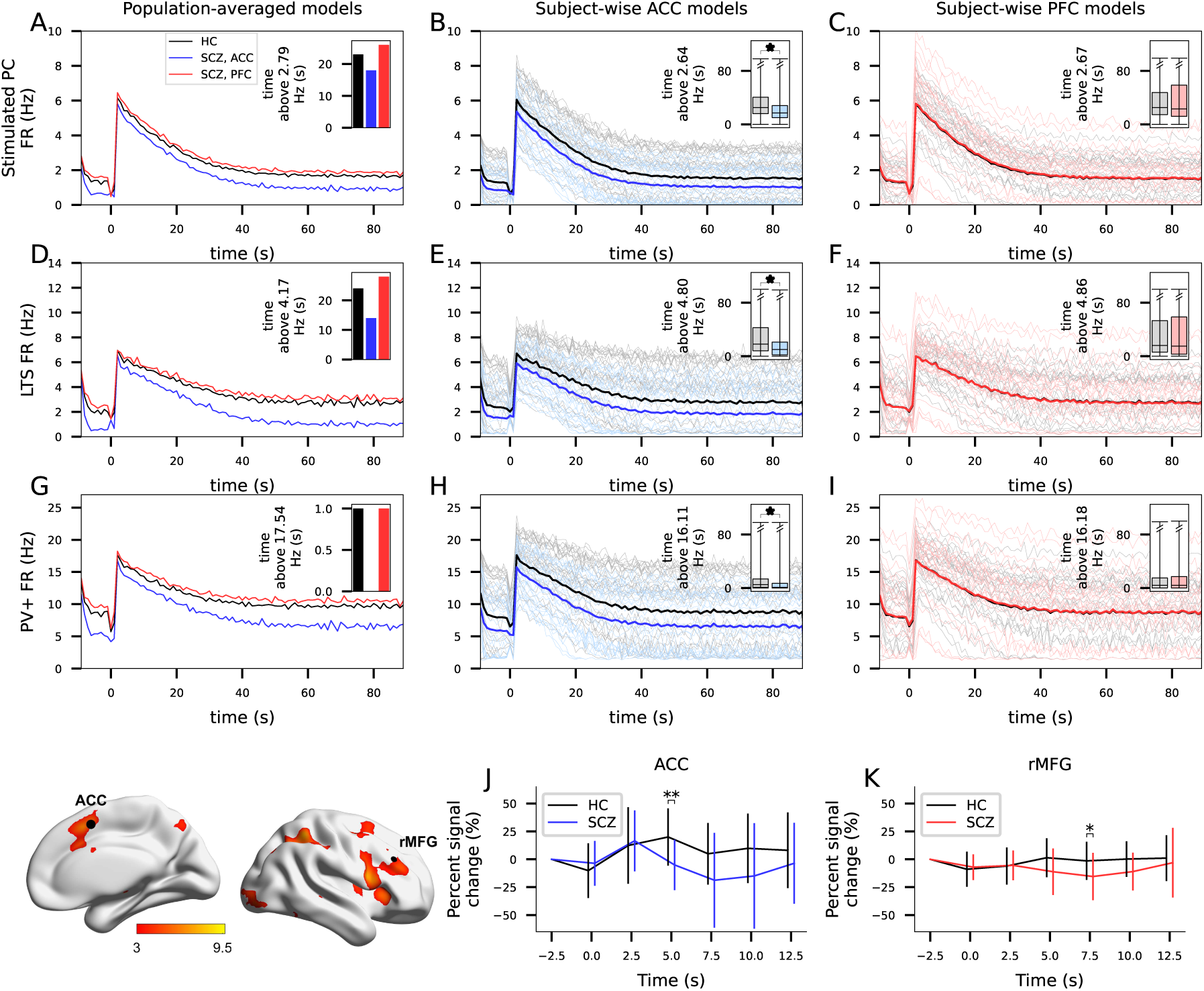
ACC-like variants of ion-channel expression in pyramidal cells decrease the predicted network response to the WM-inducing stimulus, and decreased ACC activity in a WM task in SCZ is supported by fMRI data. **A,D,G**: Average firing rate of pyramidal cells (A), parvalbumin-positive interneurons (D), and LTS interneurons (G) in the network undergoing WM-inducing stimulus from 0 s on. Black: default model. Blue, red: models where the ion-channel conductances in pyramidal cells were manipulated according to average gene expression difference of SCZ vs HC subjects in ACC (blue) or PFC (red; see Table 1). Insets show the time that the firing rate remains above 2*×* the average baseline firing rate in the default model (7.44 Hz, 6.16 Hz or 13.32 Hz in pyramidal cells, parvalbumin-positive cells and LTS interneurons, respectively). **B,E,H**: Firing rates of pyramidal cells (B), parvalbumin-positive interneurons (E), and LTS interneurons (H) in the subject-wise models according to the ACC data. The dim gray and blue curves show the firing rates of the HC and SCZ models, respectively. Both types of models were obtained by calculating subject-wise gene-expression factors (the expression level normalized by the average expression level in HCs) and multiplying the corresponding pyramidal cell ion-channel conductances by these factors. The thick curves show the averages across HCs (black) or SCZ subjects (blue). Insets: For each subject-wise model, the time above 2*×* the average baseline firing rate in HCs models (6.88 Hz, 6.66 Hz or 12.85 Hz in pyramidal cells, parvalbumin-positive cells and LTS interneurons, respectively) was determined. The bars show the mean and SD of these durations. **C,F,I**: The experiment of panels B, E, and H repeated using the expression data from PFC. **J–K**: Hemodynamic responses in ACC (J) and PFC (K) in response to target stimuli in the 2-back task — the brain maps are illustrated on the left. The curves show the mean hemodynamic responses across the subjects at time instants 0, 2.5, 5, 7.5, 10, and 12.5 s (relative to the value at −2.5 s), and the ticks represent the SD (HC: black, SCZ: blue, red). The asterisks show the significant differences between SCZ (blue, red) and HC (black; U-test, p*<* 0.05), and the double asterisks show the significant differences that survive multiple corrections (p*<* 0.05*/*12).

To test the hypothesis of reduced neural activity in the ACC of patients with SCZ during a WM task in vivo, we analysed an open fMRI dataset recorded during a N-back WM task where the participants had to view a sequence of letters and indicate whether each letter matched a predefined target (0-back) or the one presented one or two trials earlier (1-back/2-back) [Repov̌s and Barch, 2012]. In the 2-back condition with high WM load, the hemodynamic response at 5 seconds post-stimulus for target stimuli was significantly decreased in SCZ (N=15) compared to HCs (N=27) in the ACC (U-test, p=0.0029*<*0.05/12; Fig. 3J). There was also a trend of decreased hemodynamic response for target stimuli in SCZ compared to HCs in the right middle-frontal gyrus within the PFC at 7.5 seconds post-stimulus, but this was not significant when corrected for multiple comparisons (U-test, p=0.029*>*0.05/12; Fig. 3K). The hemodynamic responses for non-target stimuli were not significantly different in SCZ compared to HC (Fig. S3A–B). We did not find differences in 0-back and 1-back conditions (Fig. S3C–J).

We next analysed the predicted effects of the SCZ-associated gene-expression alterations on Ca^2+^ dynamics and spectral activity in the network. The reduction in the delay-period firing activity for the ACC-like expression variants was accompanied by decreased Ca^2+^ concentration and NMDAR-mediated currents in the pyramidal cells (Fig. 4A–B) and a decreased spectral power of pyramidal cell firing in all frequency ranges apart from the high gamma band (Fig. 4C–D, blue). The effects of the PFC-like variants on Ca^2+^ activity and spectral density were less consistent but showed an opposite, milder trend (mildly higher Ca^2+^ activity and increased spectral power of pyramidal cell firing in theta, alpha and beta frequencies; Fig. 4C–D, red).

**Figure 4:**
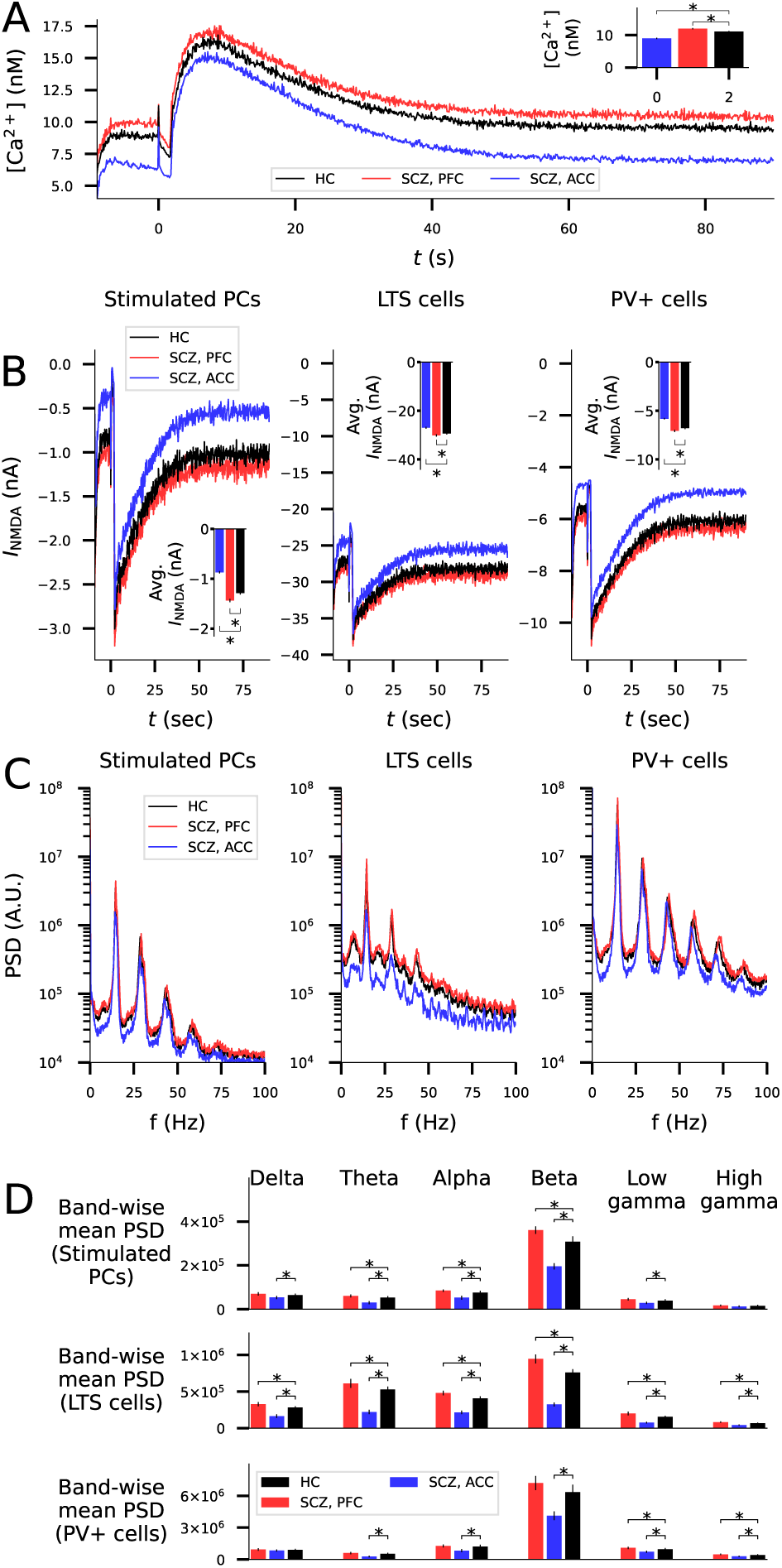
SCZ-associated alterations of expression of ion channel-encoding genes in ACC (but not PFC) decrease Ca^2+^ activity in pyramidal cells and spectral power of the firing activity in all frequency bands and all populations. **A**: The intracellular Ca^2+^ concentration in pyramidal cell somata (averaged across the 582 pyramidal cells and 5 repetitions). The WM-inducing stimulus is given at *t* = 0 sec. Inset: mean and SD of the time-averaged Ca^2+^ concentration across the 5 repetitions. **B**: The summed NMDAR-mediated currents in pyramidal cells (averaged across the 582 pyramidal cells and 5 repetitions). Inset: mean and SD of the time-averaged NMDAR-mediated currents across the 5 repetitions. **C**: The PSD calculated from the firing rate data for the stimulated pyramidal cells, LTS cells and parvalbumin-positive cells in the HC model (black) and under ion channel expression level changes typical to SCZ in the ACC (blue) and PFC (red). The fast fourier transform was used for calculating the PSD, and the PSD was downsampled by averaging 20 consecutive data points for clarity, resulting in a resolution of 0.2 Hz. **D**: The band-averaged PSD estimated from the data of (C). The bars represent the mean PSDs across delta (0.5–4 Hz), theta (4–8 Hz), alpha (8–13 Hz), beta (13–30 Hz), low gamma (30–70 Hz), and high gamma (70–100 Hz) bands. Asterisks in panel (D) and in the insets of panels (A) and (B) denote statistically significant differences between SCZ and HC when Bonferroni-corrected for multiple tests (U-test, p*<* 0.05*/*6).

The model predictions above take into account the effects of the altered expression of ion channel-encoding genes on the activity of cortical pyramidal cells but not inhibitory neurons. Similar to pyramidal neurons, cortical interneurons rank among the brain cell types most affected by expression of SCZ-associated genes [Skene et al., 2018, Ruzicka et al., 2024]. If inhibitory neurons exhibit same changes in the pattern of ion channel expression as the pyramidal neurons, these changes may compensate for the decreased excitability underlying the predicted impairment of WM response. Here, we implemented those ion-channel conductance parameter changes of Table 1 that were applicable to the parvalbumin-positive and LTS neuron models, namely, we adapted the conductance parameters *g̶*_Na,t_ and *g̶*_K,DR_ in both interneuron types according to the SCN1B and KCNB1 expression data and the *g̶*_Ca,L*−*type_ and *g̶*_Ih_ conductances in the LTS neurons according to the CACNA1C, CACNA1D, and HCN1 expression data. Our model predicts that the sustained activity is shorter-lived in SCZ compared to HC in ACC also when inhibitory neurons express the ion-channel conductance changes suggested by gene-expression data (Fig. S4A–I). In the personalized simulations, the difference between the duration of sustained activity was significant for all neuron populations in the ACC but not in the PFC (Fig. S4B–C,E–F,H–I). Likewise, the Ca^2+^ activity and NMDAR-mediated currents were significantly smaller in the pyramidal cells for the ACC-like variants even when similar conductance changes were implemented in the inhibitory neurons (Fig. S4J–K).

Our network model is based on the observation that persistent activity can be induced by strong [Ca^2+^]-elevating stimulus through an HCN channel-dependent mechanism in PFC [Winograd et al., 2008]. To test if our model predictions for the SCZ-associated expression alterations hold in brain areas lacking this mechanism, we replaced the HCN channel-mediated increase in firing rate by an external increase of AMPAR-mediated inputs whose dynamics mimicked the HCN channel-mediated increase in firing. Namely, we fitted an exponential decay form to the firing rate curve of HCs in Fig. 3A (*τ* =16 sec; Fig. S5A), and added AMPAR-mediated synaptic inputs to the stimulated pyramidal cells whose amplitude we adjusted to keep the current magnitude of the WM-induced increase in AP firing (S5B). We then repeated the simulations of Fig. 3 with this external WM-inducing mechanism. The simulations showed that the ACC-like variants decreased the predicted network activity in response to the external WM-like inputs (S5C–K).

Taken together, our model predicts that SCZ-associated alterations of expression of ion channels in the ACC, in particular those responsible for A-type or delayed-rectifier K^+^ channels or T-type Ca^2+^ currents, can lead to shortening of sustained firing caused by the WM-inducing stimulus in cortical microcircuits and a generic decrease in NMDAR-mediated currents and intracellular Ca^2+^ concentration in pyramidal cells. These predictions are supported by fMRI data from the ACC of SCZ patients and HCs collected during a WM task.

### 2.3 Plasticity pathway modelling suggests enhanced baseline synaptic conductance and impaired LTP in SCZ that correlate with reduced sustained firing in the ACC

We have previously shown by mass-action-based modelling of synaptic plasticity that the gene expression alterations in SCZ, especially those measured in the ACC, can significantly impair the postsynaptic LTP in the cortex. However, it is not known how these effects relate to the sustained firing in response to the WM-inducing stimulus and whether they can compensate for the SCZ-associated impairments of the sustained firing response. Here, we repeated the simulations of synaptic plasticity of [Mäki-Marttunen et al., 2024] and studied their association with the predicted impairments of sustained firing.

We first performed population-averaged simulations (gene-wise and combined), where the concentration of protein phosphatase A (PKA), protein phosphatase 1 (PP1), phospholipase 2 (PLA2), phosphodiesterase 4 (PDE4), protein kinase C (PKC), calcium/calmodulin-dependent kinase II (CaMKII), Na^+^/Ca^2+^ exchanger (NCX), neurogranin (Ng), Gi and Gq proteins, calbindin (Calb), and diacylglycerol kinase (DAGK) were altered based on the CommonMind RNA expression data (see Table 2). In the ACC of individuals with SCZ, the baseline conductance was predicted to be mildly increased by decreased PPP1CA and PPP1CC expression (+2.0% effect on baseline conductance) and increased PRKAR2A expression (+1.1%) but less affected by other expression alterations (effects ranging from −0.14% to +0.26%), and the combination of all expression alterations yielded a +4.3% increase in the baseline conductance (Fig. S6A, blue). Likewise in the PFC, the baseline conductance was mildly increased by decreased PRKAR2A expression (+1.4%) but was less affected (ranging from −0.2% to +0.2%) by other changes in expression levels: the combination yielded a similar +1.4% increase in the baseline conductance (Fig. S6A, red). Qualitatively similar effects were observed in the baseline concentration of membrane-bound GluR1 receptors (Fig. S6B), a major contributor to the total synaptic conductance in the model [Mäki-Marttunen et al., 2020, Mäki-Marttunen et al., 2024].

**Table 2:**
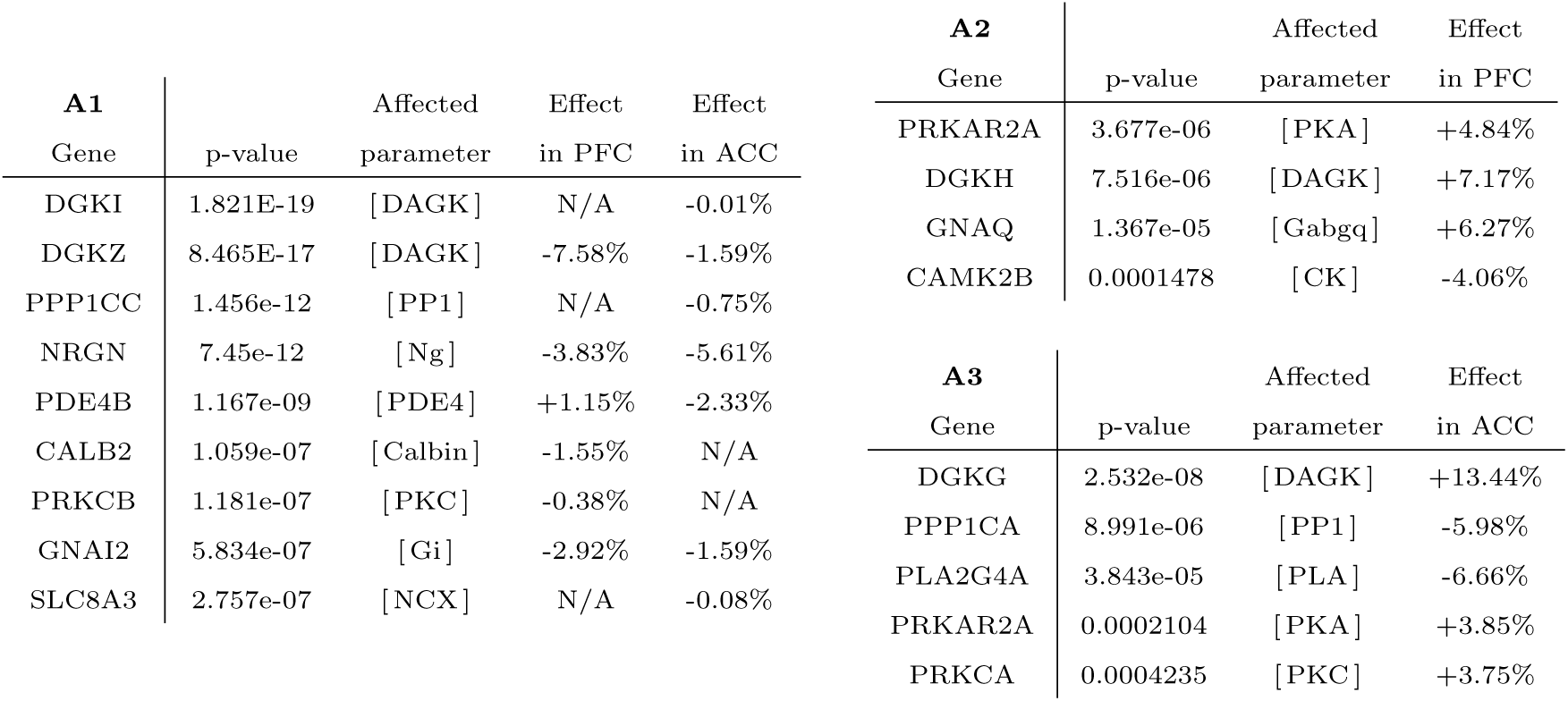
Table of plasticity-regulating genes associated with SCZ and the alterations in expression level in SCZ compared to controls according to post-mortem expression data in PFC and ACC. **A**: Genes that included SNPs moderately or highly associated (p-value *<* 5*×*10*^−^*^6^) with the risk of SCZ according to GWAS data [Trubetskoy et al., 2022]. **B–C**: Genes whose post-mortem expression in the PFC (B) or ACC (C) was moderately or highly associated (p-value *<* 5*×*10*^−^*^5^) with SCZ according to the CommonMind data [Hoffman et al., 2019]. See Table 1 for details.

We tested the effects of these parameter changes on the plasticity response to an high-frequency (16 Hz) stimulation lasting 100 sec, which can lead to a short-lived synaptic depression followed by an LTP lasting for more than 30 min (Fig. S6C–Q), as analyzed in [Mäki-Marttunen et al., 2024]. Decreased PPP1CA/PPP1CC expression had little effect on synaptic conductance at 30 min post-stimulus, but the increase in PRKAR2A expression mildly increased the synaptic conductance at the long time scale (Fig. S6C) similar to its effect on the baseline synaptic conductance. In addition, the decreased expression of PLA2G4A in the ACC decreased the synaptic conductance at 30 min post-stimulus, and the increased expression of PRKCA in the ACC had an opposite effect (Fig. S6C). The combination of all expression alterations yielded a modest 0.4% (ACC) or 0.1% (PFC) decrease in the absolute synaptic conductance at 30 min after stimulus onset (Fig. S6C). However, given the excitatory effects on the baseline conductance, the potentiation at 30 min after the stimulus onset was weakened from +125% to +115% (ACC) or +122% (PFC; Fig. S6D).

To test the significance of the observed effects, we performed subject-wise simulations where a personalised model was adjusted for each subject in the RNA expression data set based on individual expression data and the results of the SCZ simulations were compared to those of the HC simulations. The baseline synaptic conductance was significantly increased in SCZ according to both ACC (37.3 pS in SCZ, 35.6 pS in HC, p=4.3*×*10*^−^*^12^; U-test) and PFC (35.7 pS in SCZ, 35.5 pS in HC, p=0.0077; U-test) data. The weakening of both the potentiation at 30 min post-stimulus was also significant in both ACC and PFC (ACC: LTP amplitude +97% in SCZ and +103% in HC, p=4.3*×*10*^−^*^5^; PFC: LTP amplitude +103% in SCZ and +109% in HC, p=0.0063; U-test; Fig. 5A–B).

**Figure 5:**
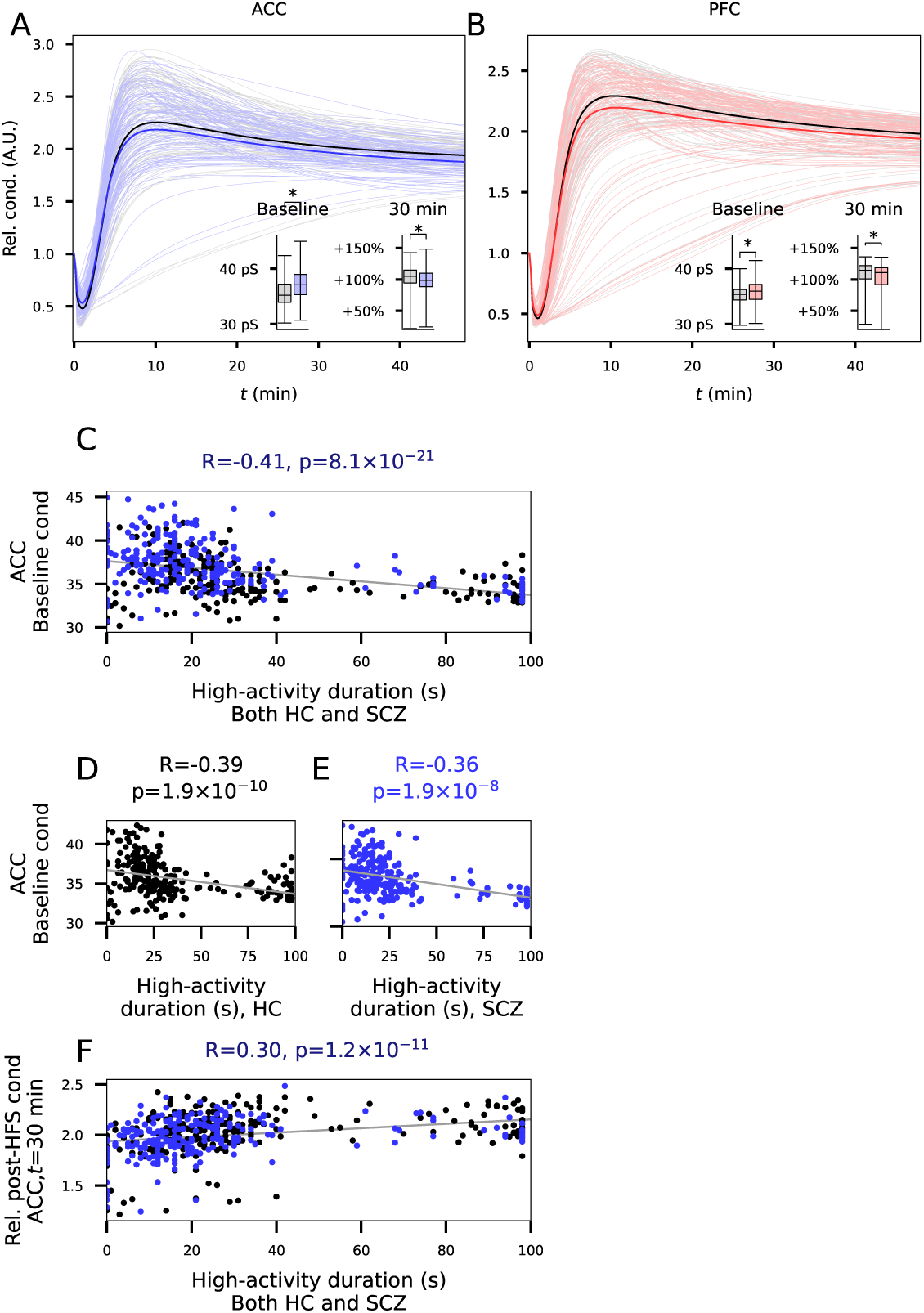
SCZ-associated alterations in the expression of plasticity-regulating genes in ACC lead to impaired potentiation at the long time scale and an increased baseline synaptic strength that correlate with decreased sustained firing. **A,B**: Time courses of relative synaptic conductance (synaptic conductance at different time instants normalized by baseline conductance). The dim curves represent models where the initial concentrations of the proteins of Table 2 were adjusted based on individual expression data from the CommonMind data set in ACC (A) or PFC (B) — the gray curves represent the HC subjects while the blue (A) and red (B) curves represent the SCZ subjects. The thick curves represent the averages across the subject-wise simulations. Left inset: Baseline conductances. Right inset: Relative synaptic conductance 30 min after stimulus onset. **C**: The baseline conductance for SCZ (blue) and HC (black) subjects plotted against the predicted duration of sustained activity following the WM-inducing stimulus from the same subjects. The correlation coefficient (R) and the p-value of the association test printed above the graph. **D–E**: The data of (C) printed separately for HC (D) and SCZ (E) subjects. The association of the predicted LTP amplitude and the duration of the sustained firing is qualitatively similar in HC (D) and SCZ (E) subjects compared to the pooled sample (C). **F**: The predicted amplitudes of the potentiation at 30 min after stimulus onset, i.e., the synaptic conductance at *t* =30 min normalized by baseline conductance, plotted against the predicted duration of sustained activity following the WM-inducing stimulus from the same subjects.

We next tested the association between the predicted response to the WM-inducing stimulus in the network model and the predicted synaptic conductance at baseline and after plasticity-inducing stimulus. In the ACC, the predicted baseline synaptic conductance was significantly anti-correlated with the duration of the sustained firing after a WM-inducing stimulus both in the whole population (r=-0.41, p=8.1*×*10*^−^*^21^, t-test; Fig. 5C) and in the HC and SCZ populations separately (r=-0.39, p=1.9*×*10*^−^*^10^ in HC; r=-0.36, p=1.9*×*10*^−^*^8^ in SCZ; Fig. 5D–E). Consistent with the results from Fig. S6, the duration of the sustained firing after a WM-inducing stimulus was also positively correlated (r=0.30, p=1.2*×*10*^−^*^11^) with the amplitude of the LTP at 30 min post-stimulus onset (Fig. 5F). In the PFC, by contrast, the duration of the sustained firing after a WM-inducing stimulus was associated with a larger baseline synaptic conductance (r=0.52, p=6.1*×*10*^−^*^31^) but not with LTP amplitude (r=0.00, p=0.97; Fig. S7A–D).

Although the models of [Neymotin et al., 2016] and [Mäki-Marttunen et al., 2024] cannot be combined due to the differences in spatiotemporal scales, it is possible to test whether the SCZ-associated increase in baseline synaptic conductance predicted by the model of [Mäki-Marttunen et al., 2024] could compensate for the SCZ-associated decreased sustained firing predicted by the model of [Neymotin et al., 2016]. To do this, we adapted the baseline synaptic conductance of the excitatory-to-excitatory synapses in the model of Fig. 3. We did this both in population-averaged models (Fig. 3A,D,G) where the synaptic conductance was 4.3% (ACC) or 1.4% (PFC) larger in SCZ than HC and in the personalized models (Fig. 3B–C,E–F,H–I) where the synaptic conductance was adapted in a subject-wise manner based on the data from Fig. 5A–B. Our simulations suggest that duration of the sustained firing following WM-inducing stimulus is still significantly shorter in SCZ compared to HC according to the models tuned to gene expression data in the ACC even when taking into account the predicted increase in the baseline conductance of excitatory to excitatory synapses (Fig. S8).

Taken together, our analyses suggest that altered expression of genes associated with synaptic plasticity via the PKA and PKC pathways as measured post-mortem in the ACC, and to a lesser extent in the PFC, leads to an increase in baseline synaptic conductance and an impairment of LTP. These effects were associated with shorter-lived sustained network firing.

### 2.4 Genotype-phenotype analysis supports a role in shaping WM performance for genes that encode ion channels and regulate plasticity

To validate our findings with genetic and behavioural WM data, we used cognitive data from a TOP study where 619 participants with SCZ and 992 HCs performed an LNS WM task and were genotyped [Haatveit et al., 2023]. Similar to [Mäki-Marttunen et al., 2024], we determined SCZ PRSs focusing on ion channel encoding genes or plasticity-regulating genes, in addition to traditional genome-wide PRS of SCZ for each participant (see Methods). We tested the association between the different PRSs and WM test performance.

We fit a linear model for the WM test performance against age and sex and the SCZ PRSs (one PRS at the time). The PRS based on the full genome was not associated with WM performance (Fig. 6A, green), but the PRSs based on ion channel-encoding genes (Fig. 6A; blue), synaptic plasticity-regulating genes (Fig. 6B; red), or both (Fig. 6A; magenta) were associated with poorer WM performance. When only Na^+^ and HCN channel encoding genes or K^+^ channel encoding genes were included in the PRS, there was no association with the WM performance (Fig. 6A; blue with vertical or horizontal stripes), but there was an association between poorer WM performance and the PRS based on Ca^2+^ channel encoding genes (Fig. 6A; blue, diagonal stripes). Poorer WM performance was also associated with the PRS strictly based on the 11 ion channel-encoding genes included in the computational analysis, listed in Table 1 (Fig. 6A; blue with gray dots).

**Figure 6:**
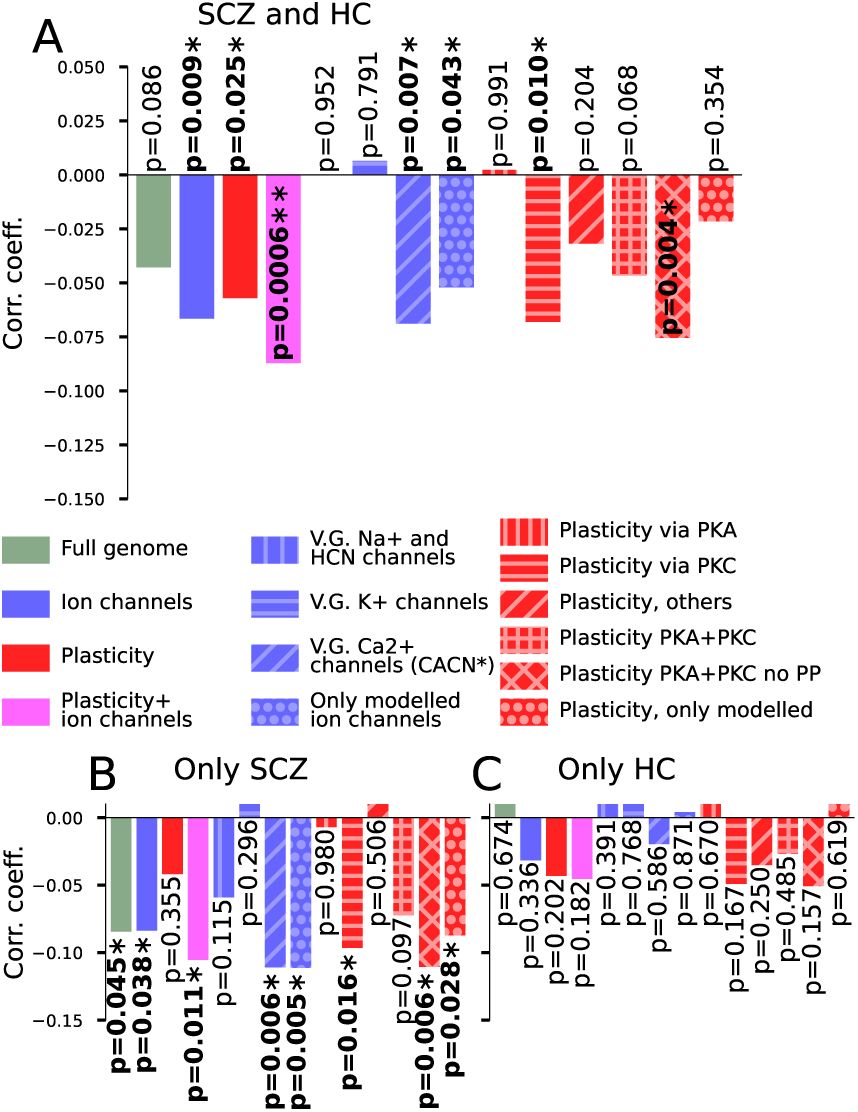
Genotype-phenotype analysis validates the model-based findings. **A**: Correlation coefficients (sample size N=1611) between the WM performance and eight different SCZ PRSs, namely, the PRSs based on SNPs in 1) all human genes (gray), 2) genes encoding ion channels (as defined in Table S2 from [Mäki-Marttunen et al., 2024]; blue), 3) plasticity-regulating genes (as defined in Table S2 from [Mäki-Marttunen et al., 2024]; red), 4) plasticity-regulating genes and genes encoding ion channels (i.e., the union of (2) and (3); magenta), 5) genes encoding Na^+^ or HCN channels (a subset of (2); blue with vertical stripes), 6) genes encoding K^+^ channels (a subset of (2); blue with horizontal stripes), 7) genes encoding Ca^2+^ channels (a subset of (2); blue with diagonal stripes), 8) genes of Table 1 (blue with dots), 9) genes regulating plasticity primarily through the PKA pathway ([Mäki-Marttunen et al., 2024]; red with vertical stripes), 10) genes regulating plasticity primarily through the PKC pathway ([Mäki-Marttunen et al., 2024]; red with horizontal stripes), 11) genes regulating plasticity primarily through other pathways ([Mäki-Marttunen et al., 2024]; red with diagonal stripes), 12) genes regulating plasticity primarily through PKA or PKC pathways (red with horizontal-vertical grid), 13) genes regulating plasticity primarily through PKA or PKC pathways excluding protein phosphatase-encoding genes (red with diagonal grid), or 14) genes of Table 2 (red with dots). For all these PRSs, SNPs that had a p-value smaller than 1 *×* 10*^−^*^8^ according to the SCZ GWAS data [Trubetskoy et al., 2022] were included. The bars marked with asterisks indicate statistically significant associations in a linear model with age and sex as covariates (t-test, p*<*0.05), and the double asterisks indicate significance when Bonferroni-corrected for multiple tests (p*<*0.05/14). **B–C**: The analysis of (A) repeated for SCZ (B; N=619) or HC (C; N=992) samples separately.

We next dissected the PRS based on plasticity-regulating genes. When we categorized the plasticity-regulating genes into genes acting through the PKA or PKC pathway, or other plasticity-related genes (see Table S2 in [Mäki-Marttunen et al., 2024]), the PRS based on PKC-pathway genes was associated with poorer WM performance (Fig. 6A; red with horizontal stripes) whereas those based on PKA-pathway (Fig. 6A; red with vertical stripes) and other plasticity-regulating genes (Fig. 6A; red with diagonal stripes) were not. A PRS based on plasticity-regulating genes acting through PKA or PKC pathway was not associated with WM performance (Fig. 6A; red with gray grid), but a PRS based on a smaller set of genes where protein-phosphatase encoding genes were left out was associated with poorer WM performance (significant also when Bonferroni-corrected for multiple tests, p*<*0.05/14; Fig. 6A; red with diagonal grid). The PRS based only on the modelled plasticity-regulating genes (Table 2) was, however, not associated with the WM performance (Fig. 6A; red with gray dots). To show the robustness of our genotype-phenotype findings, we performed additional unplanned association analyses.

Most of the above associations were also significant when only SCZ patients (Fig. 6B) were included, and importantly, both PRSs based on modelled genes only were associated with poorer WM performance (Fig. 6B; blue and red bars with gray dots). The analysis based on HC data only did not show significant associations (Fig. 6C). The findings of Fig. 6A were replicated when a stricter (p-value for SCZ diagnosis *<* 1 *×* 10*^−^*^10^) SNP limit was used (apart from the association of the PRS based on the 11 modelled ion channel-encoding genes; Fig. S9A) and when a more relaxed (*<* 1 *×* 10*^−^*^6^) limit was used (Fig. S9B). Our findings were also robust when IQ was added as a covariate (Fig. S9C), when only age was used as a covariate (Fig. S9D), and when both covariates (age, and sex) were ignored (Fig. S9E). Finally, most of the associations were replicated when we used PRSs calculated from a bipolar disorder (BD) GWAS [O’Connell et al., 2025] (Fig. S9F), which suggests that WM impairments may be associated with ion-channel and plasticity-related genetic risk for psychotic disorders in general rather than SCZ specifically.

The associations between WM performance and the different PRSs were dependent on the cohort. In the group of subjects that performed the WAIS-III LNS test, all associations apart from the association of poorer WM performance and the PRS based on plasticity-regulating genes acting through the PKC pathway remained significant and the correlation coefficients between the PRSs and the WM performance was more negative than in the pooled data (correlation coefficients ranged between −0.087 and 0.006 in the pooled sample, Fig 6A, and between −0.112 and −0.009 in the WAIS-III sample, Fig S9G). By contrast, in the group of subjects that performed the MCCB LNS test, none of the associations reached a level of significance (Fig. S9H).

Taken together, the genetic and behavioural data suggest that the polygenic risk of SCZ mediated by SNPs in ion channel subunit encoding genes and genes regulating synaptic plasticity is associated with poorer WM test performance. In particular, the negative association between the SCZ PRS based on SNPs in the genes whose expression data were used for modelling (Table 1) and the WM test performance partially validates our model predictions of impaired delay-period firing in SCZ.

### 2.5 Single-gene analyses suggest a role for CACNA1I overexpression in increasing SCZ risk and decreasing WM performance

The association between SCZ PRSs and experimentally measured WM performance (Fig. 6) suggests that the 11 genes included in our simulations (Table 1) may contribute to impairments in WM performance. However, it remains unclear whether the same or different genes among the 11 are important in the simulations versus the PRS-WM performance associations. In addition, genetic associations do not necessarily act through genes in proximity to the variant, as risk-associated variants may influence the expression of other genes rather than their host gene. To address these questions, we (1) repeated the analysis shown in Fig. 6 using single-gene risk scores for all modelled ion channel-encoding genes, and (2) performed a two-sample MR analysis using GWAS summary statistics for SCZ and cis-eQTL data from brain tissue [De Klein et al., 2023] to identify putative causal relationships between ion channel gene expression and SCZ.

We first formed single-gene risk scores for the genes of Table 1 and determined their association with the WM test performance. Apart from KCND3, KCNJ6, KCNMA1, KCNQ3, and SCN1B that had no SNPs with SCZ-risk p-value less than 1 *×* 10*^−^*^8^, the other genes had 115 (CACNA1C), 80 (HCN1), 70 (CACNA1I), 3 (CACNA1D), 2 (KCNB1), or 1 (ATP2A2) SNPs passing the p-value threshold. The single-gene risk score of CACNA1I was associated with poorer WM performance (p=0.045*<*0.05), with the risk-scores of CACNA1C and CACNA1D showing non-significant effects in the same direction (Fig. 7A).

**Figure 7:**
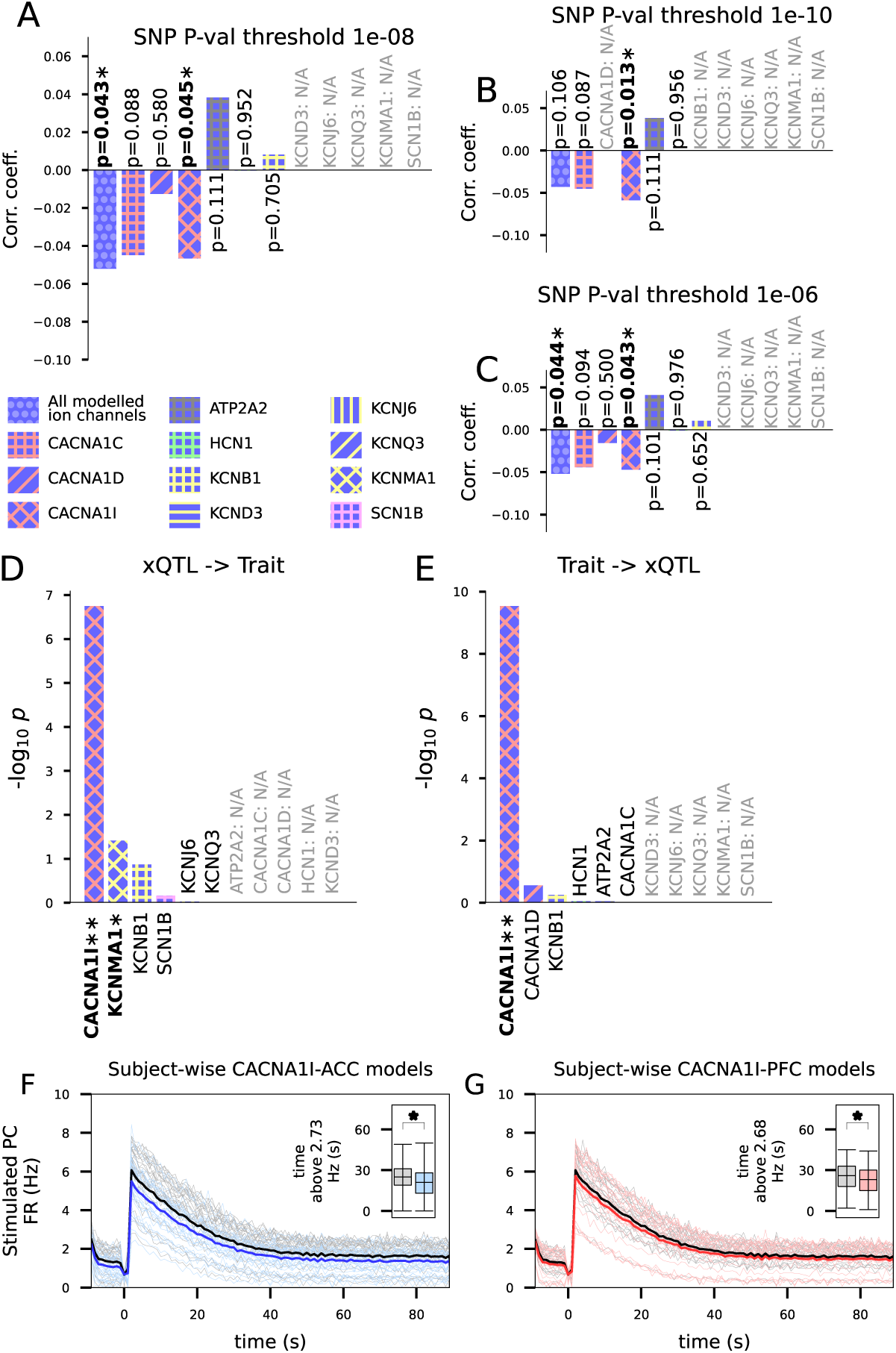
The role of CACNA1I in WM and SCZ risk is supported by multimodal analyses. **A–C**: The experiment of Fig. 6A (association of WM test performance with PRSs) repeated for single-gene risk scores using a SNP p-value threshold of 1 *×* 10*^−^*^8^ (A), 5 *×* 10*^−^*^10^ (B) or 5 *×* 10*^−^*^6^ (C). The y-axis shows the correlation coefficient between the summed SCZ risk in the SNPs within one gene (ATP2A2, CACNA1C, CACNA1D, CACNA1I, HCN1, KCNB1, KCNQ3, or KCNMA1) and the WM test performance (sample size N=1611). None of the associations were significant when Bonferroni-corrected for multiple tests (p*<*0.05/11). **D–E**: The significance of different genes in the MR approach where the predicted gene expression was treated as the exposure and SCZ liability as the outcome (D) or vice versa (E). The y-axis shows the negative of the 10-base logarithm of the p-value. The bars marked with asterisks indicate statistically significant associations (Wald z-test, p*<*0.05), and double asterisks indicate a significance also when Bonferroni-corrected for multiple tests (p*<*0.05/11). The expression of the genes labeled as N/A were not successfully predicted. **F–G**: Firing rates of stimulated pyramidal cells in response to the WM-inducing stimulus in networks where the T-type Ca^2+^ channel conductance was scaled according to subject-wise CACNA1I expression data from ACC (F) or PFC (G). See Fig. 3B–C for details.

In the MR analyses using gene expression from cortical brain tissue, we observed support for causal associations with CACNA1I. The genetic liability to greater expression of CACNA1I was associated with increased SCZ risk (*β*=0.23, SE = 0.04, p = 1.78*×*10*^−^*^7^; Fig. 7D). Genetic liability to increased KCNMA1 expression was associated with reduced risk of SCZ, but this association did not survive correction (*β*=-0.07, SE = 0.03, p = 0.039; Fig. 7D). In the reverse direction, genetically proxied SCZ liability was associated with increased expression of CACNA1I (*β*=3.53, SE = 0.56, p = 2.91*×*10*^−^*^10^; Fig. 7E). No evidence was observed for the other ion channel-encoding genes examined (ATP2A2, CACNA1C, CACNA1D, KCNB1, KCND3, KCNJ6, KCNQ3, SCN1B; Fig. 7D–E).

To confirm the relevance of increased CACNA1I expression alone for delay-period activity, we repeated the simulations of Fig. 3 such that we only adapted the LVA Ca^2+^ channel conductance according to the gene-expression data. Similar to the polygenic simulations of Fig. 3, the SCZ population exhibited shorter response to the WM-inducing stimulus when tuned with the CACNA1I expression data from the ACC (p=4.2*×*10*^−^*^6^; U-test; Fig. 7F). The corresponding simulations for CACNA1I expression data from the PFC also showed significantly shorter response (p=0.032; U-test; Fig. 7G) but with smaller effect size (the average duration of the sustained firing in the pyramidal cells was 10.0% shorter in SCZ simulations for the PFC but 18.2% shorter for ACC; Fig. 7F–G).

Taken together, these results suggest that 1) SCZ risk within the CACNA1I gene is associated with poorer WM test performance, 2) increased CACNA1I expression in brain tissue has a putative bidirectional causal relationship with SCZ risk, and 3) increased CACNA1I expression alone as measured in the ACC of SCZ patients predicts a compromised delay-period activity. These observations support our interpretation that compromised WM test performance in patients with SCZ is at least partly mediated by increased expression of CACNA1I.

## 3 Discussion

Our work combined four data modalities (post-mortem gene expression, behavioural WM test, GWAS, and fMRI) to study the mechanisms behind WM deficits in SCZ. We linked these data through three different methodologies as follows (Fig. 1). First, we (i) inferred the effects of differential expression in SCZ (post-mortem RNA expression from ACC and PFC used) on delay-period activity and LTP dynamics using *biophysically and biochemically detailed modelling*. We found that the SCZ-associated differential expression of KCND3, CACNA1I, and KCNB1 in ACC predicted a shorter response to the WM-inducing stimulus (Figs. 3A–I, S2) and that of PPP1CA, PPP1CC, and PLA2G4A predicted an increase in baseline synaptic strength and a decrease in LTP amplitude (Figs. 5, S6) that correlated with the shorter response to the WM-inducing stimulus (Fig. 5). These predictions suggest a mechanism for the observation of decreased BOLD signal activity in the ACC during WM tasks in SCZ (Figs. 3J, S3). Next, we (ii) calculated *SCZ PRSs based on gene sets of interest* and determined their association with behavioural WM test performance. We found that high PRSs based on ion channel-encoding or plasticity-regulating genes were significantly associated with poorer letter-number sequencing (LNS) test performance (Fig. 6). When we narrowed down the gene base of the PRSs, this association remained significant when we only used the set of genes applied in the biophysically detailed modelling (Tab. 1), and even when we used a single-gene risk score that only included the SNPs within the gene CACNA1I (Figs. 6–7). Finally, we (iii) studied the relationship between gene expression and SCZ risk by applying *Mendelian randomization (MR) to SCZ GWAS summary statistics and cis-eQTL data from brain tissue*. We found a putative bidirectional causal link between increased CACNA1I expression and the risk of SCZ. In particular, our study presents a triangulation of evidence to suggest the role of CACNA1I SNPs in mediating both the SCZ risk and an impaired WM test (LNS) performance. Our simulations suggest that this happens through increased CACNA1I expression leading to increased SK currents that decrease the pyramidal cell firing activity and response to WM-inducing stimulation.

Within the ACC, our model predicts that overexpression of KCND3, CACNA1I, and KCNB1 — especially in combination — produces a broad-band reduction in firing activity across neuron populations. This reduced excitability is particularly relevant in light of theoretical accounts on the role of ACC. [Shenhav et al., 2013] suggest that the ACC has a central role in evaluating the expected value of cognitive control. In the setting of a WM task such as LNS or N-back task, this could be understood as evaluating the need for processing information associated with the past stimuli based on the new stimulus — or alternatively the need for processing the new stimulus based on the synaptic inputs encoding the past stimulus. In this framework, the predicted decrease in ACC activity on the time scale of seconds (Fig. 3) suggests that the computation of cognitive control demands and the evaluation of stimulus history may be compromised in SCZ, potentially leading to less efficient allocation of control during WM tasks. Another theory posits that ACC participates in cognitive control by detecting conflicts between competing representations of the stimuli and their transformations while the PFC resolves such conflicts [Carter and Van Veen, 2007]. In the framework of this theory, our model predictions of decreased ACC activity suggest that SCZ patients be more likely to confuse different representations of the stimuli or suboptimally transform these representations.

In the PFC, CACNA1I expression was also mildly increased, and single-gene model variants using PFC CACNA1I data showed significantly reduced persistent firing during the delay period (Fig. S2F, 7G), consistent with a detrimental effect on WM maintenance. However, in the full PFC model, overexpression of HCN1 partially compensated for this and other gene-expression changes, resulting in mildly increased firing during the delay period (Fig. S2) and significantly elevated spectral power in theta, alpha, beta, and low-gamma bands across all simulated cell types (Fig. 4D). This pattern is difficult to map directly to ion-channel-level mechanisms because HCN currents have been linked to both impairment and improvement of WM, depending on the neuromodulatory context [Ramos et al., 2006, Wang et al., 2007, Wu et al., 2024, Thuault et al., 2013]. Computational modelling provides an efficient means of elucidating complex ion-channel-activation relationships such as these. Previous computational studies show that both pro-excitation and shunting inhibition effects of HCN are possible, depending on the stimulus strength and interaction with other ion channels [George et al., 2009, Kase and Imoto, 2012, Migliore and Migliore, 2012, Mäki-Marttunen and Mäki-Marttunen, 2022] — yet, in the present modelling framework, the excitatory consequences dominate.

Our predictions of suppressed ACC activity were robust to model variations, including parallel genetic effects in interneurons (Fig. S4) and compensation of baseline synaptic strengths (Fig. S8), and they are partly supported by human imaging and electrophysiological evidence. Our own BOLD analysis based on an open dataset (Fig. 3J) and previous work [Kerns et al., 2005] indicate altered ACC dynamics in SCZ, and another study shows evidence for group*×*time interactions although not for uniform hypo-activation [Becerril and Barch, 2013]. At the oscillatory level, SCZ is associated with widespread EEG abnormalities both during WM tasks [Lynn and Sponheim, 2016] and at rest [Hunt et al., 2017, Perrottelli et al., 2021], and our model’s broad-band reduction in ACC firing aligns with reports of reduced evoked theta, alpha, and beta activity during WM encoding in early- and adult-onset SCZ [Haenschel et al., 2009, Pachou et al., 2008, Wang et al., 2026]. While experimental findings suggest alterations also in the gamma band [Haenschel et al., 2009, Cho et al., 2006, Minzenberg et al., 2010], our model only showed significant SCZ–HC gamma-band differences in the two classes of interneurons.

Combining gene expression data with our biochemically detailed synapse model suggests that, in ACC of SCZ patients, baseline synaptic conductance is increased whereas LTP amplitude is reduced (Fig. 5). This prediction can help explain the decreased within-test learning that has been observed in patients in addition to an overall impaired test performance [I‧mamŏglu et al., 2025]. Interestingly, the predicted changes in LTP amplitude and increased baseline synaptic conductance correlated with decreased network firing. This supports the hypothesis that downregulation of excitability via SCZ-associated ion-channel genes is accompanied by compensatory changes in PKA/PKC-pathway gene expression that increase baseline synaptic strength, or conversely that altered synaptic signalling drives changes in intrinsic excitability. Such coordinated expression changes are consistent with activity-dependent transcription underlying homeostatic plasticity [Dubes et al., 2019], in which synaptic scaling in response to altered neuronal activity is at least partly mediated by transcriptional regulation of synaptic genes.

Our behavioural-genetic analyses partly support an involvement of SCZ risk in ion channel-encoding and plasticity-related genes, including CACNA1I, in compromising WM performance. In the pooled sample and specifically in participants who completed the WAIS-III LNS test, but not the MCCB version, higher SCZ genetic risk and CACNA1I single-gene risk were associated with poorer LNS performance, and these associations were significant only among SCZ patients when analysed separately. The lack of association in the MCCB sample could possibly be due to relatively smaller SCZ group (N=248, compared to N=371 patients that performed the WAIS-III LNS test) or differences in the details of the two versions of the LNS test. The observation that the association was only significant in the SCZ group is consistent with models proposing that SCZ risk variants contribute to subtle distributed effects on synaptic and network function while measurable effects on cognition and connectivity arise primarily in the presence of environmental risk factors [van Os and Kapur, 2009, Schmitt et al., 2023]. Thus, while these associations warrant testing in larger and more diverse samples and with multiple WM paradigms, the genetic results provide partial human-level support for the mechanisms suggested by our simulations.

It is important to emphasize that our computational framework is a generic neocortical model, not specifically fitted to PFC or ACC microcircuit data. The HCN-mediated upregulation of pyramidal excitability following WM-inducing stimuli is grounded in PFC electrophysiology [Winograd et al., 2008], whereas comparable mechanisms in ACC have not been established, motivating our alternative ACC model with WM-related external AMPAR input (Fig. S5). In this variant, SCZ-tuned ACC models show reduced responsiveness to WM-associated input, consistent with our main findings. Nevertheless, additional region-specific differences in cellular properties, connectivity, and neuromodulation between ACC and PFC are expected but were beyond the scope of the present work.

RNA expression data used for our simulation experiments may be affected not only by genetic and environmental risk factors but also by phenomena directly or indirectly caused by the disorder. Antipsychotic use is one of the main such confounders for which there is no data in the CommonMind dataset. However, apart from mood stabilizers potentially affecting KCNQ3 and SCN1B expression [Smolin et al., 2012], we did not find evidence of use of antipsychotics or homeostatic regulation affecting the RNA expression for the genes we included in our modelling framework. For example, a recent macaque study [Schulmann et al., 2023] found RNA expression of many genes affected by antipsychotic use (4–56 genes using a p-value threshold of 5*·*10*^−^*^6^, depending on the type and dose of antipsychotics), but this gene set did not overlap with our modelled genes (Tables 1, 2). Nevertheless, we cannot fully exclude treatment or homeostatic compensation as contributors to the observed expression patterns.

Taken together, our simulations and genetic analyses suggest that SCZ risk variants in cell-membrane ion-channel and synaptic plasticity pathways contribute to altered ACC and PFC excitability, synaptic strength, and oscillatory dynamics that impair WM performance. These mechanistic insights can guide future work that stratifies SCZ and HC cohorts into genetically informed subgroups, enabling more precise hypotheses about subgroup-specific WM deficits.

## 4 Methods

### 4.1 Biophysically detailed model of HCN-channel mediated persistent activity in a cortical circuitry for representation of WM

We used the model of [Neymotin et al., 2016] for simulating the circuit response to the WM-inducing stimulus and neural activity during the delay period. The model consists of 582 five-compartment pyramidal neurons, each consisting of a soma, basal dendrite, and three consecutive apical dendritic sections, 97 single-compartment parvalbumin-positive interneurons, and 97 single-compartment low-threshold spiking (LTS) interneurons (Fig. 8A). All populations were distributed into layers 2, 5 and 6 and were recurrently connected to each other in a layer-specific manner [Neymotin et al., 2016]. All cell types included the passive current, transient Na^+^ currents and delayed-rectifier K^+^ currents. In addition, the pyramidal cells included the following ion channels: L-, N- and T-type Ca^2+^ channels, HCN channels, M-, A- and D-type K^+^ channels, and BK-like Ca^2+^-activated K^+^ channels, and the LTS interneurons included the L-type Ca^2+^ channels, HCN channels and SK channels (Fig. 8A). LTS and pyramidal neurons include the description of Ca^2+^ dynamics needed for the Ca^2+^-activated K^+^ channels. In pyramidal cells, also HCN channels are modulated by Ca^2+^through an intracellular signalling network consisting of mGluR and IP_3_R activation and the subsequent release of Ca^2+^ from the ER into the cytosol (Fig. 8B).

**Figure 8:**
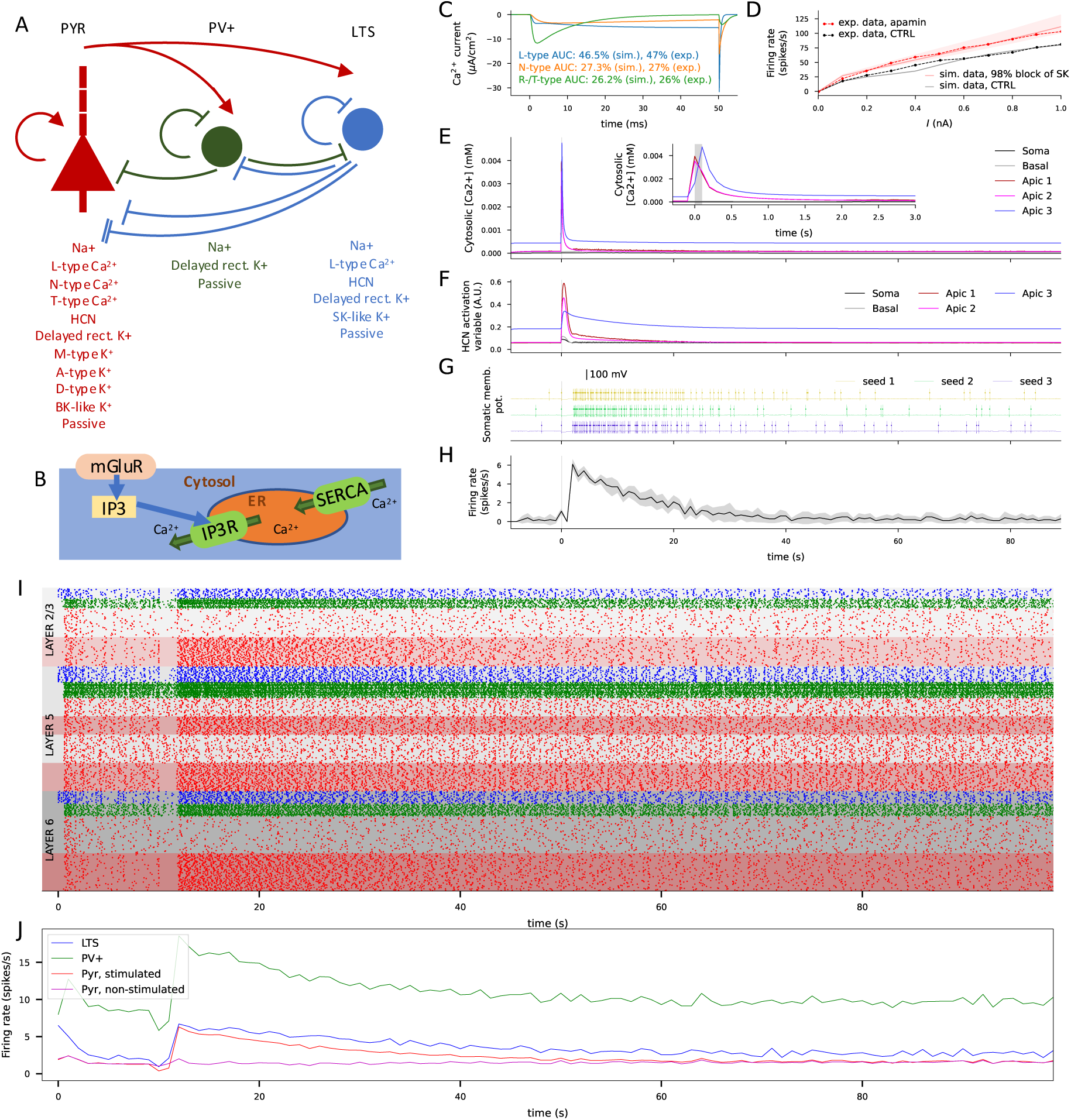
Illustration of the model components and model behaviour. **A**: The neuron types and their structural properties and connectivity. The pyramidal neurons (red) consisted of the soma, a basal dendritic compartment and three apical dendritic compartments, each expressing some of the active ion channels (see [Neymotin et al., 2016]). The parvalbumin-positive interneurons (green) and LTS interneurons (blue) consisted of a single compartment expressing active Na^+^ and K^+^ channels and a passive leak channel, and the LTS neuron model also included an HCN current, Ca^2+^ currents and a Ca^2+^-activated K^+^ current. All neuron types projected to all other neuron types in a layer-specific way (see [Neymotin et al., 2016]). The parvalbumin-positive and LTS neurons projected GABAAR-mediated currents to other neurons (denoted by green and blue inhibitory connections, respectively), but the LTS neurons additionally projected GABABR-mediated currents to pyramidal neurons (denoted by a blue double inhibitory connection). All compartments with Ca^2+^ channels and Ca^2+^-activated K^+^ channels include a model for intracellular Ca^2+^ concentration that is increased by incoming Ca^2+^ currents and decays toward resting-state Ca^2+^ concentration in the absence of inputs. **B**: The pyramidal neuron dendrites also include a model describing mGluR-, IP3R- and SERCA-pump-mediated Ca^2+^ dynamics. In this model, inputs to the mGluR activate the IP3R, which leads to a Ca^2+^ release from the ER to the cytosol. In addition to Ca^2+^ extrusion to the extracellular medium, Ca^2+^ is sequestered to the ER by the SERCA pump. **C**: The simulated L-, N-, and T-type Ca^2+^ currents in response to a 50-ms somatic voltage clamp to 0 mV (from −110 mV). The fractions of each current of the total AUC (between 0 and 50 ms) in the simulated data (“sim.”) fit well to the experimentally measured (“exp.”; [Almog et al., 2009]) fractions. **D**: The simulated (solid) and experimentally measured (dashed; [Guan et al., 2015]) f-I curves, i.e., firing frequencies of the neuron in response to different somatic DC amplitudes, in the absence (black) and presence (red) of SK channel blockade. The dim red area represents apamin efficiencies 95–100% (98% displayed with the solid curve). **E**: The cytosolic Ca^2+^ concentration in different compartments of the pyramidal neuron in response to the WM-inducing stimulus in a single-neuron simulation. **F**: The activation variable of the HCN current in different compartments of the pyramidal neuron in response to the WM-inducing stimulus in a single-neuron simulation. **G**: The somatic membrane potential of the pyramidal neuron in response to the WM-inducing stimulus in a single-neuron simulation, repeated three times with different random number seeds affecting the timing of the background synaptic firing. The long-lasting increase in firing in response to the WM-inducing stimulus is due to the increase in HCN current activity [Neymotin et al., 2016]. **H**: The firing rate of a pyramidal neuron as a function of time in a single-neuron simulation, determined from the three repeated simulations using a bin size of 1 sec. The curves show the mean and the shaded area represents the SD of the firing rate. **I**: The population spike train of the full network model in response to the WM-inducing stimulus at *t*=10 sec. The lower half of each pyramidal neuron population represents the randomly picked neurons that received the WM-inducing stimulation while the upper half represents the non-stimulated neurons. **J**: The average firing rate in each population of the network. The pyramidal neuron firing rates were divided into two signals, one for the stimulated half (red) and one for the non-stimulated one (magenta).

The WM-inducing stimulus is modeled using the persistent firing-inducing stimulation protocol of [Neymotin et al., 2016]. In this protocol, AMPARs and NMDARs of all neurons and the mGluRs of pyramidal cells are stimulated for 100 ms. This leads to a long-lasting activation of the HCN channels, giving rise to increased excitability of the pyramidal neurons in the time scale of seconds [Neymotin et al., 2016].

We made a few adjustments to the model of [Neymotin et al., 2016] to align it with single-cell electrophysiology data on the activity of the small-conductance Ca^2+^-activated SK channels that mediate the medium afterhyperpolarization period as well as Ca^2+^ channels in pyramidal cells. Namely, we aimed to adjust the model to fit the data of [Almog et al., 2009], where it was estimated that 29% of the charge transfer in Ca^2+^ currents was mediated by L-type channels, 17% by N-type channels, 16% by R-type channels (LVA/MVA), 17% by P-type channels, and 21% by Q-type channels. Since our model did not contain P- or Q-type channels and described the T-type channels instead of R-type channels, we used the relative proportion of L-, N-, and R-type Ca^2+^ channel currents from their summed contribution (47%, 27%, and 26%) as the target Ca^2+^-channel balance. We also aimed at reproduction of the effects of SK channel blockade on pyramidal cell firing activity in rat somatosensory cortex [Guan et al., 2015]. To achieve this, we first replaced the SK channel model used in [Neymotin et al., 2016] and [Shah et al., 2008] where the conductance depended on both the intracellular [Ca^2+^] and membrane potential by the SK channel model used in [Hay et al., 2011] where it depended only on the intracellular [Ca^2+^], using the same maximal conductance (0.0442 S/cm^2^) as in the soma in the model of [Hay et al., 2011]. By hand-tuning and the use of the bisection method, we then adjusted the L-type, N-type, and T-type Ca^2+^ channel conductances to 120%, 30%, and 84% of their respective values in [Neymotin et al., 2016]. We also set the somatic Ca^2+^ channel conductances to 25% of the corresponding conductances in the apical dendrite (10% in [Neymotin et al., 2016]). In L5 pyramidal cells, we also adopted a hot zone of Ca^2+^ channels by multiplying the L- and T-type Ca^2+^ channel conductances in the most distal apical dendritic compartment by 100 [Hay et al., 2011]. These adjustments gave a good fit to the data of [Almog et al., 2009] (Fig. 8C), and it also fit well to the data of [Guan et al., 2015] where blockade of SK channels by apamin increased the L5 pyramidal cell firing rate in the rat somatosensory cortex (Fig. 8D). Qualitatively similar results were obtained with the model lacking the hot zone of Ca^2+^ channels (Fig. S10). We used this pyramidal cell model henceforth, and included the hot zone in all layer 5 pyramidal cells but not layer 2/3 or 6 pyramidal cells.

Similar to the single-cell pyramidal cell model of [Neymotin et al., 2016], a strong synaptic activation of the pyramidal cells, driven by AMPAR, NMDAR and mGluR activations that model a WM-inducing stimulus, leads to IP_3_R-mediated increase in intracellular Ca^2+^(Fig. 8E). This leads to increased HCN-channel activation (Fig. 8F) and augmented firing rate (Fig. 8G–H). The same HCN-channel-mediated mechanism underlies the increased firing rate during the delay period in the full network model (Fig. 8I–J) — in the network simulations, we adjusted the synaptic weights (*w*_I*→*E_ from 0.1 to 0.05, *w*_E*→*parvalbumin*−*positive_ from 0.002 to 0.008, *w*_E*→*LTS_ from 0.002 to 0.004, and *w*_E*→*E_ from 0.00025 to 0.0005, see [Neymotin et al., 2016]) to retain realistic AP rates.

#### 4.1.1 Modelling SCZ-associated alterations of ion channel properties

We used the CommonMind data set [Hoffman et al., 2019] of post-mortem gene expression in HCs and SCZ patients. We followed the approach of [Mäki-Marttunen et al., 2024] by selecting the genes of interest as the intersection of genes whose products were included in our computational model and genes that are likely to be expressed in neocortex. We also applied the same method as in [Mäki-Marttunen et al., 2024] to determine the expected change in ion-channel gene expression in SCZ patients compared to HCs. In brief, we used the DESeq2 normalization of the RNA expression of ion-channel or ion-channel-subunit-encoding genes in PFC and ACC from 426 or 478 subjects (215 HC and 211 SCZ samples for PFC, and 251 HC and 227 SCZ samples for ACC) and imputed their neuronal expression. For the imputation, we used CIBERSORTx [Steen et al., 2020] and a reference single-cell RNA dataset [Zhang et al., 2016] to determine the RNA expression in neurons. Most of the ion channel-encoding genes were successfully imputed but for some there were too few reads available, which caused the imputation to fail — these genes were ignored in the present work.

We identified the genes that were differentially expressed in SCZ patients compared to HCs using a linear model where age, sex and PMI were controlled for. We used a cut-off p-value of 5*×*10*^−^*^4^ — this resulted in 4 genes in the PFC data set and 6 genes in the ACC data set (Table 1). We also included GWAS hit genes that had a p-value smaller than 5*×*10*^−^*^6^ in the data of [Trubetskoy et al., 2022] (Table 1). However, we left out genes that did not show single-cell-type specificity for excitatory neurons (in categories Cell type enhanced or Group enriched) in the Human Protein Atlas (https://www.proteinatlas.org) to avoid basing our model predictions on altered expression in genes that are unlikely to be expressed in pyramidal neurons. In the population-averaged simulations, we used the exponentiated beta coefficient as a measure of how much more (*>* 1) or less (*<* 1) expression of the gene is there to be expected in SCZ compared to HC. In the subject-wise simulations, we calculated for each individual the relative expression of each gene normalized by the average expression of the gene among HCs. The simulations were then run for each individual separately, and group differences were tested for the model output, namely, the duration of the high-frequency firing of pyramidal cells after a WM-inducing stimulus. Our main analysis was centered on the effects of gene expression differences in pyramidal cells because their genetics and ion channel properties are generally better mapped, but we also tested the robustness of our model predictions to similar changes in interneurons.

#### 4.1.2 Mapping of ionic currents and genes in the computational model

The genes identified using the above method are listed in Table 1. Of these, ATP2A2 is a SERCA pump isoform, whereas all other genes (CACNA1C, CACNA1D, CACNA1I, HCN1, KCNB1, KCND3, KCNJ6, KCNMA1, KCNQ3, and SCN1B) encode ion channels or their subunits. Increased ATP2A2 expression leads to an accelerated uptake of Ca^2+^ from the cytosol to the ER in the pyramidal cells (see Fig. 8). CACNA1C and CACNA1D contribute to the L-type Ca^2+^ channel-mediated currents, whereas CACNA1I contributes to the T-type Ca^2+^ channel-mediated current, and SCN1B encodes the beta subunit of a Na^+^ channel mediating fast Na^+^ currents. As for the K^+^ channel-related genes, KCNB1 encodes the Kv2.1 subunit mediating a delayed rectifier K^+^ current, KCND3 encodes the shal-related Kv4.3 subunit mediating an A-type K^+^ current, KCNJ6 encodes the GIRK2 subunit mediating a GABA_B_R-activated current, KCNMA1 encodes a BK-channel subunit mediating the Ca^2+^ and voltage-dependent large-conductance K^+^ currents, and KCNQ3 encodes the Kv7.3 subunit mediating an M-type K^+^ current. In each case, we determined the model parameters for the SCZ case by multiplying the corresponding maximal conductance parameter by the ratio of gene expression in SCZ patients normalized by that in HCs (shown in Table 1). When altering the L-type Ca^2+^ channel conductance, we used the average gene-expression ratio of CACNA1C and CACNA1D as the factor for the maximal conductance.

The effects of altered expression of some of the key risk genes are difficult to predict due to the computational model being incomplete at the genetic level. This applies to SCN1B and KCND3 in particular. In pyramidal cells, the proteins encoded by these genes are likely to be expressed alongside other proteins in the family, and thus changes in the concentration of these ion channels may lead to smaller effects on the underlying ion-channel current in the model than expected by the 1:1 relationship. Here, we used genetic and electrophysiological data to make a conservative estimate of the effects of altered expression of these genes. For SCN1B, electrophysiological experiments [Hull et al., 2020] showed that a homozygous knock-out of the gene caused a 38.4% decrease in the peak current density in cortical pyramidal cells. Similar data were observed for KCND3: A homozygous knock-out of KCND3 decreased the *I_A_* peak density by 45.4% in cortical pyramidal neurons [Norris and Nerbonne, 2010]. These observations are in line with tissue-level RNA expression data from the Human Protein Atlas indicating that SCN1B accounts for 52% of the total RNA expression of Na^+^-channel *β* subunits in excitatory neurons (SCN1B, SCN2B, SCN3B and SCN4B had expressions of 541.7, 95.9, 164.8 and 248.5 nTPM, respectively) and that KCND3 accounts for 48% of the total RNA expression of A-type K^+^ channel subunit-encoding genes (KCND3, KCND2 and KCNA4 had expressions of 56.9, 33.7 and 27.7 nTPM, respectively; Human Protein Atlas, https://www.proteinatlas.org/, referenced Aug 19th 2025). We thus estimated that 40% of both current species in the model were contributed to by these respective genes and thus the effects of all gene-expression alterations on the channel conductances were respective to this ratio (shown in parentheses in Table 1).

### 4.2 Biochemically detailed model of post-synaptic plasticity in the cortex

We used the recent model of synaptic plasticity mediated by the PKA, PKC, and CaMKII pathways [Mäki-Marttunen et al., 2024] for simulating the effects of altered expression of plasticity-associated genes on the synaptic conductance and its development during LTP/LTD-inducing stimulus protocols. The model consists of a single post-synaptic compartment (spine) including 256 molecular species and 365 mass-action-based reactions between them [Mäki-Marttunen et al., 2024]. The system is activated by 3-ms Ca^2+^ (120 particles/ms) and neuromodulatory (5 particles/ms norepinephrine, 10 particles/ms glutamate, and 10 particles/ms acetylcholine) inputs that are repeated for 100 times at a frequency of 16 Hz, and the synaptic conductance can be estimated from the model output using a statistical model of AMPAR tetramer formation [Mäki-Marttunen et al., 2020]. Following the approach of [Mäki-Marttunen et al., 2024], we adjusted the initial concentration of some of the proteins to model the effects of altered gene expression in a similar fashion as done for the genes of Table 1 (Table 2). In each simulation, the stimulus started at 6.6 hours (biological time) to ensure a steady state. We quantified the numbers of GluR1 and GluR2 subunits at the synapse membrane for each simulation. For GluR1, we determined the numbers of both S831-phosphorylated and non-S831-phosphorylated subunits. We then calculated the number of AMPAR tetramers residing at the membrane and the probabilities of the tetramers being of homomeric GluR2-type, homomeric GluR1-type with and without S831-phosphorylated subunits, or GluR1-GluR2 heteromers, and estimated the expected total synaptic AMPAR conductance [Mäki-Marttunen et al., 2020], which was used as the output of the model.

### 4.3 BOLD-fMRI data analysis

The dataset of HCs and participants with SCZ was obtained from OpenNeuro (https://openneuro.org/datasets/ds000115/versions/0 [Repov̌s and Barch, 2012]. The task data consisted of the N-back task, a WM task where participants are presented with a sequence of letters in each block and have to assess whether the shown letter is the same as a pre-specified target letter (0-back) or the same as a previously shown (1-back/2-back) letter in the sequence [Repov̌s and Barch, 2012]. All scans were acquired on a 3T Tim TRIO scanner [Repov̌s and Barch, 2012]. We included participants older than 18 years old, and excluded participants without task design information (subject 72) or with reduced number of volumes (subject 81, 1-back). For the current analyses, we did not include the data from siblings of participants with SCZ. The final sample sizes were of N=27 HCs and N=15 participants with SCZ. For each participant, data consisted of a T1-weighted structural images (TR = 2400 ms, TE= 3.16 ms, FOV = 256 mm, flip angle=8*^◦^*, voxel size = 1 mm x 1 mm x 1 mm) and three task-related functional T2* images (TR = 2500 ms, TE= 27 ms, FOV = 256 mm, flip angle = 90*^◦^*, voxel size = 4 mm x 4 mm x 4 mm). Each functional run consisted of two blocks of either the 0-back, 1-back, or 2-back task, resulting in three 4D volumes for each participant. Each functional scan consisted of 137 volumes. Preprocessing of the BOLD data was done using the standard pipeline of fmriprep [Esteban et al., 2019].

The pipeline included removal of the first 5 volumes to allow for stabilization of magnetic field, slice timing, realignment, artifact correction using AROMA tool, and normalization to MNI space. The functional images were submitted to first level analysis using SPM toolbox (https://www.fil.ion.ucl.ac.uk/spm/). The canonical hemodynamic response function with its temporal derivatives were used to model each event. We then calculated the contrasts to obtain target-related activation (the hemodynamic response) during the 0-back, 1-back and 2-back conditions. Extraction of percent signal change time courses for each group was done using the marsbar toolbox [Brett et al., 2002]. The regions of interest (ROIs) were defined as 5 mm spheres centred on the coordinates of maximum activation in the anterior cingulate gyrus (−2,10,50) and dorsolateral prefrontal cortex (40,30,30) as reported in a meta-analysis of the N-back task [Wang et al., 2019].

### 4.4 Experimental WM test and genetics data

We carried out post-hoc analysis for the data from the LNS experiments included in the TOP sample [Haatveit et al., 2023]. The data consisted of 1) N=371 participants with SCZ (210 male, 161 female), age 30*±*9.7 years and N=390 HCs (196 male, 194 female), age 34*±*9.7 years, who performed the WAIS-III LNS task [wec, 2003]; and 2) N=248 participants with SCZ (144 male, 104 female), age 30*±*9.1 years and N=604 (324 male, 280 female) HCs, age 33*±*8.9 years, who performed the MCCB LNS task [Nuechterlein et al., 2008]. The IQ of each participant was measured using the Wechsler Abbreviated Scale of Intelligence [Wechsler, 2011]. The WM test scores were z-transformed using the performance of the HCs as reference in both versions of the LNS task. Two HCs had performed both LNS tests and were thus included in both datasets (1 male, 1 female) — when considering the pooled sample, the average of the two z-transformed LNS scores was used as the WM performance score for these participants. An analysis of the covariates showed that WM test performance was strongly associated with IQ (correlation coefficient r = 0.51, p = 1.9*×*10*^−^*^108^) and mildly with age (r = 0.07, p = 0.003) but not with sex (p = 0.61 *>* 0.05; group difference tested by U-test). Participants with SCZ had a worse performance than HCs (p = 2.0*×^−^*^56^ *<*0.05, U-test).

All participants underwent genotyping, and PRSs of SCZ with different genomic specificity were calculated for each subject. Fourteen different gene set-based PRSs were calculated using beta coefficients and corresponding p-values from SCZ GWAS summary statistics [Trubetskoy et al., 2022] with TOP samples excluded. The largest set of genes included all genes genome-wide while other sets were based on genes encoding ion channel subunits or plasticity-regulating proteins. For each of these sets, unless otherwise stated we included SNPs with SCZ-risk p-value *<* 10*^−^*^4^ and minor allele frequency *>* 0.01. No SNP pruning was performed.

### 4.5 Mendelian randomization analyses

To test for putative causal relationships between the expression of ion channel genes and SCZ, we performed Mendelian randomization (MR). Here we performed bidirectional MR such that one analysis used gene expression as the exposure and SCZ as the outcome and the second analysis reversed the exposure and outcome. We focused our analyses on the genes used in the biophysically detailed network modelling, namely, the genes of Table 1. This result provides information on (i) if the genetic liability to ion channel gene expression is causally associated with SCZ and (ii) if the genetic liability to SCZ is causally associated with alterations in gene expression. For these analyses, we used the latest GWAS of SCZ from the Psychiatric Genomics Consortium [Trubetskoy et al., 2022] and cis-eQTL data from MetaBrain, the largest available data set of its kind, which includes cortical brain samples from 6518 participants for 18397 genes [De Klein et al., 2023]. Analyses were conducted using either the Wald ratio or inverse variance weighted approach in the TwoSampleMR R package [Hemani et al., 2018]. In line with previous work, if an instrumental variable is not present in the outcome data, we used a proxy [Namba et al., 2022, Parker et al., 2026]. Proxy variants were selected as the SNP in the highest linkage disequilibrium (LD) with the instrumented exposure genetic variant with a minimum R2 of 0.8. To estimate LD proxies, we used the European ancestry subsample of the 1000 genomes reference and PLINK v1.9 [Chang et al., 2015]. We performed correction for multiple comparisons across all MR analyses using the Benjamini-Hochberg method.

### 4.6 Statistical tests

For group differences between SCZ and HC subjects, we used the Wilcoxon rank-sum test (nonparametric; two-sided; normal approximation). For linear models (e.g., PRS-WM analyses), coefficients were tested using two-sided t-tests with degrees of freedom *df* = *n − p* where *n* was the sample size and *p* was the number of fitted variables, including the intercept and the PRS (i.e., *p* = 4 when the default covariates age and sex were used). As a robustness check, we recomputed p-values using HC3 heteroscedasticity-consistent standard errors; all conclusions were unchanged.

### 4.7 Simulation software and code accessibility

Our simulation scripts (Python, NEURON) are available at ModelDB (https://modeldb.science/2041373, password “wm”) and github (https://github.com/tuomomm/wm).

## Acknowledgements

Resources from UNINETT Sigma2 (project NN9529K/NS9529K) and CSC Finland (project 2003397) were used for high-performance simulations. Funding: Academy of Finland (330776, 336376, 318879), NIH (R01DC012947, R01DC019979, R01MH134118-01), and ARL Cooperative Agreement W911NF2220143.

## Supplementary figures and tables

**Figure S1:**
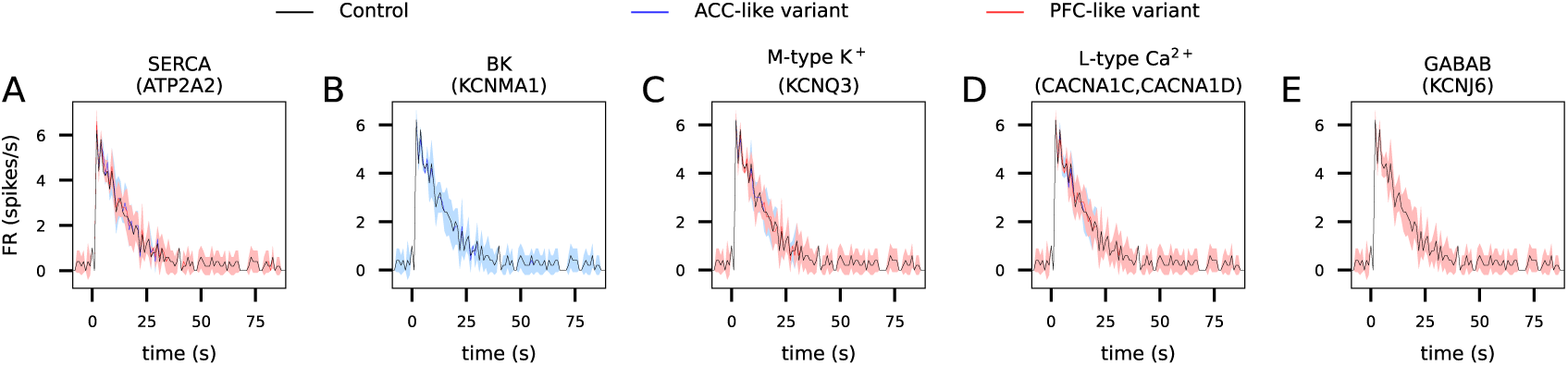
Expression-level changes of ATP2A2, KCNMA1, KCNQ3, CACNA1C, CACNA1D, and KCNJ6 associated with SCZ had little effect on the response to a WM-inducing stimulus. **A–E**: The single-neuron FR response to a WM-inducing stimulus (see Fig. 2A–E) was simulated under alterations of a single SCZ-associated ion channel as suggested by post-mortem expression data from PFC (red) and/or ACC (blue). The rate of the SERCA pump removing the Ca^2+^ from the cytosol (A), the conductance of large-conductance Ca^2+^-activated BK channel (B), M-type K^+^ channel (C), L-type Ca^2+^ channel (D), or the GABA_B_R-activated GIRK channel (E) was altered according to the expression data of the underlying SCZ-associated genes.

**Figure S2:**
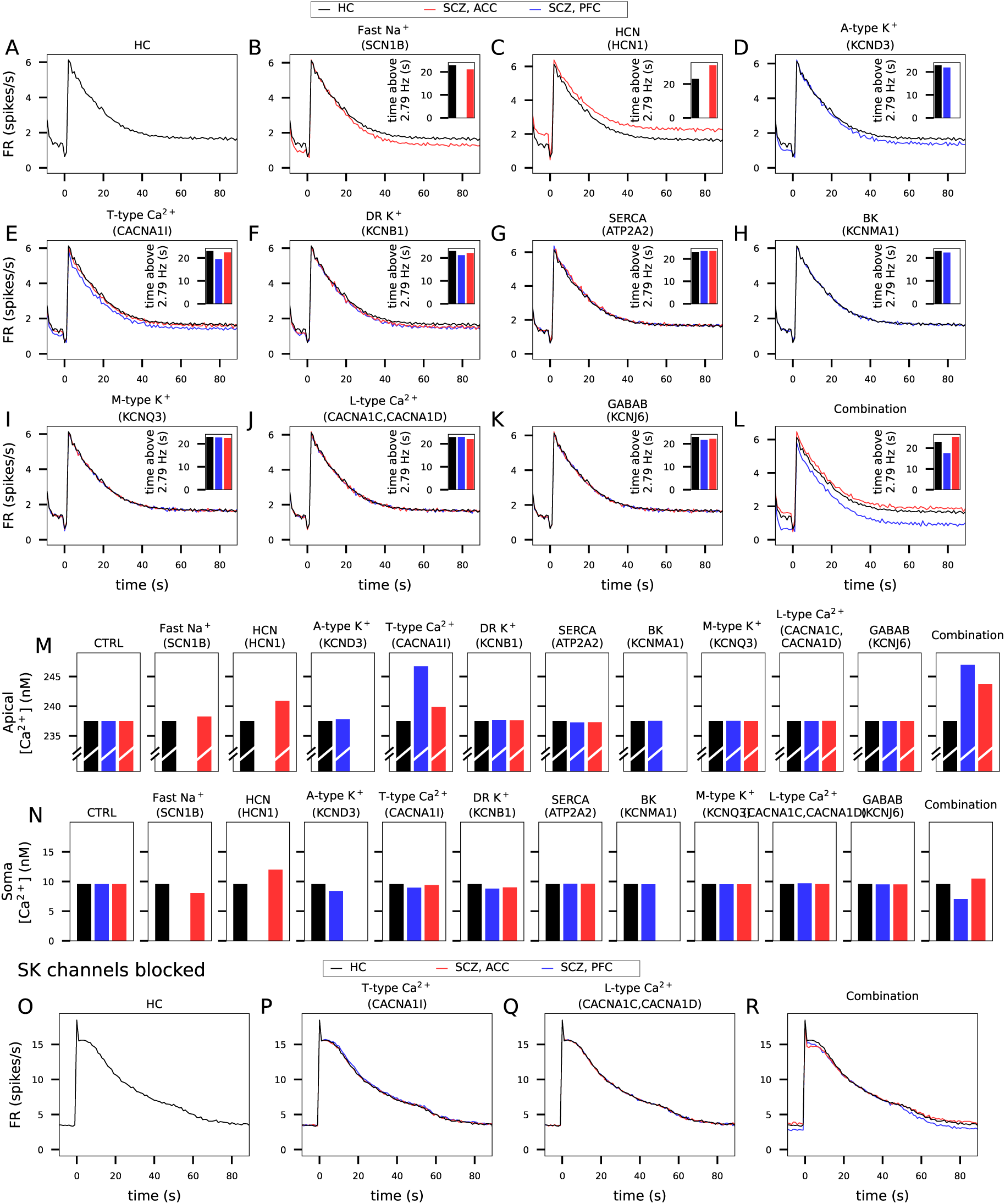
Altered expression of SCN1B, HCN1 and KCND3 had large effects on sustained network activity and that of CACNA1I had a moderate effect on the network response to the WM-inducing stimulus, while other variants had little effect, and the inhibitory effects of CACNA1I overexpression were mediated by SK currents. **A**: The firing rate of pyramidal cells in the default model. **B–K**: The effects of the population-averaged alterations (as measured in the ACC (blue) or PFC (red)) of expression of SCN1B (B), HCN1 (C), KCND3 (D), KCNB1 (E), CACNA1I (F), ATP2A2 (G), KCNMA1 (H), KCNQ3 (I), CACNA1C+CACNA1D (J), and KCNJ6 (K) on the firing activity caused by the WM-inducing stimulus. **L**: The effects of combined variants of Fig. 2K replotted for reference. **M–N**: The baseline Ca^2+^ concentrations (measured 0–3 s before the WM-inducing stimulus) in the furthermost apical dendrite (M) and in the soma (N) in the simulations of panels (A)–(L). **O**: The firing rate of pyramidal cells in the model where SK channels were blocked but no other ion-channel alterations were present. **P–Q**: The firing rate of pyramidal cells in the model where SK channels were blocked and the T-type (P) or L-type (Q) Ca^2+^ channel conductances were upscaled according to CACNA1I (P) or CACNA1C and CACNA1D (Q) expression data in SCZ. **R**: The firing rate of pyramidal cells in the model where SK channels were blocked and the combinations of ion-channel expression alterations were present.

**Figure S3:**
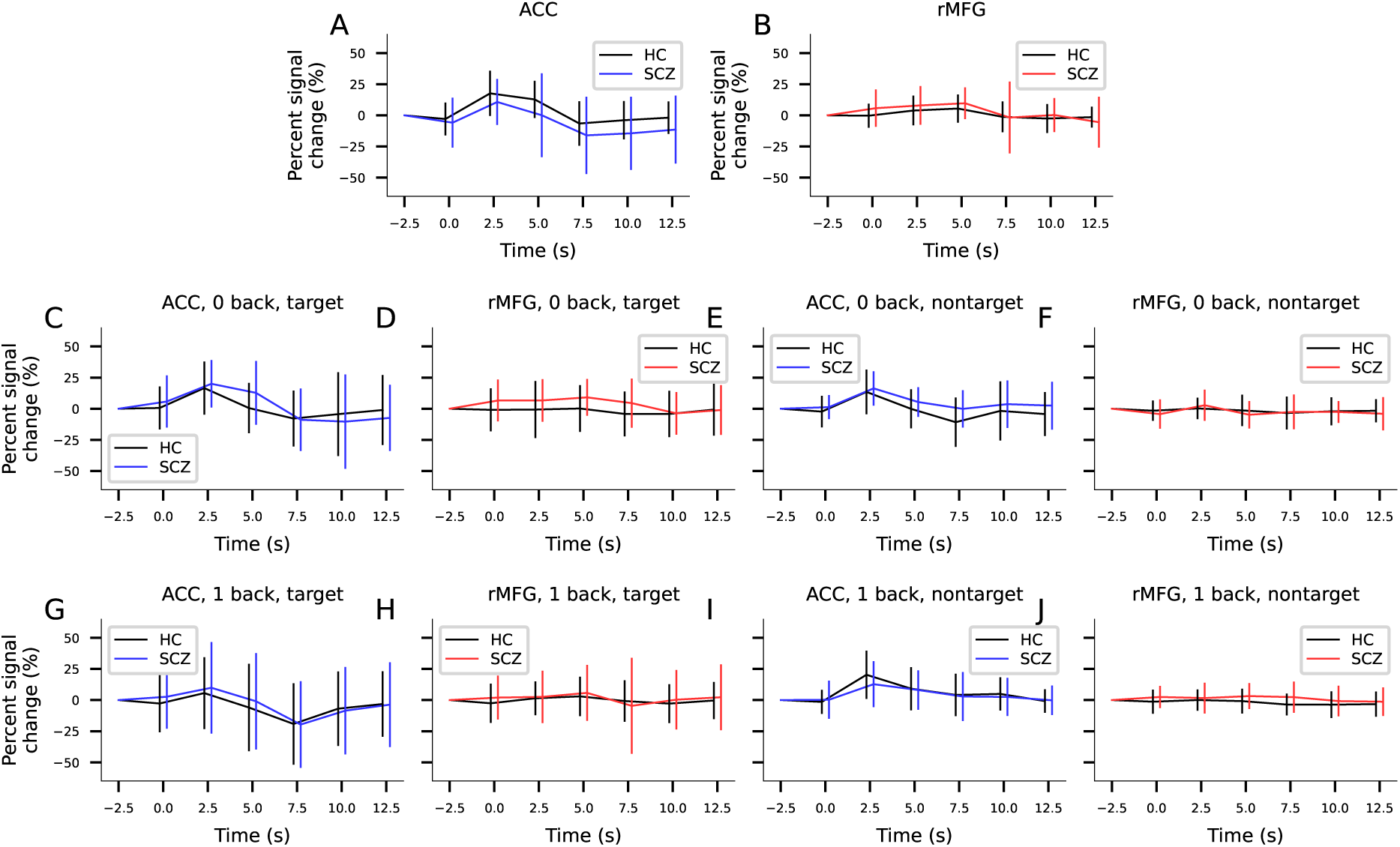
The hemodynamic responses to nontargets in the 2-back task and the responses to targets or nontargets are not significantly different between participants with SCZ and HCs. **A–B**: Hemodynamic responses in ACC (A) and PFC (B) in response to *non*target stimuli in the 2-back task. **C–F**: Hemodynamic responses in ACC (C,E) and PFC (D,F) in response to target (C–D) or nontarget (E–F) stimuli in the 0-back task. **G–J**: Hemodynamic responses in ACC (G,I) and PFC (H,J) in response to target (G–H) or nontarget (I–J) stimuli in the 1-back task. See Fig. 3J–K for details.

**Figure S4:**
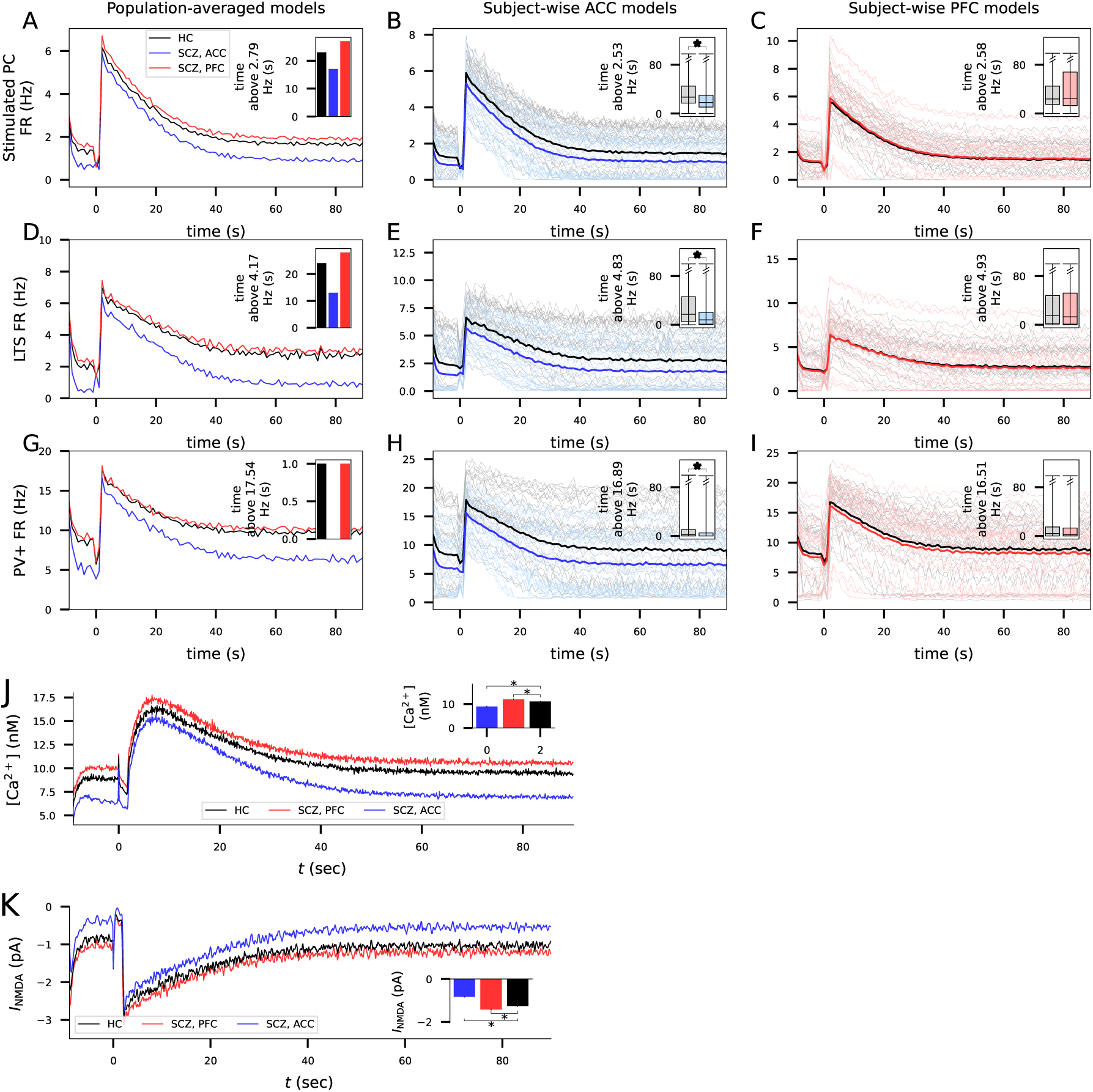
ACC-like variants of ion-channel expression in pyramidal cells *and interneurons* decrease the network response to the WM-inducing stimulus. **A–I**: The simulations of Fig. 3 were repeated so that the SCZ-associated parameter changes affected also the inhibitory neurons. Namely, the KCNB1 and SCN1B expression data affected the *g̶*_K,DR_ and *g̶*_Na,t_ parameters in all neuron populations and the CACNA1C, CACNA1D, and HCN1 expression data affected the *g̶*_Ca,L*−*type_ and *g̶*_Ih_ parameters in both pyramidal cells and LTS neurons. In the simulations of Fig. 3, in contrast, these parameter changes were only applied to the pyramidal cell population. See Fig. 3 for details. **J–K**: The simulations of Fig. 4A–B were repeated so that the SCZ-associated parameter changes affected also the inhibitory neurons. The Ca^2+^ concentration (J) and the NMDAR-mediated currents (K) in the pyramidal cells were smaller under ACC-like expression-level alterations and slightly larger under PFC-like expression-level alterations compared to HC. See Fig. 4A–B for details.

**Figure S5:**
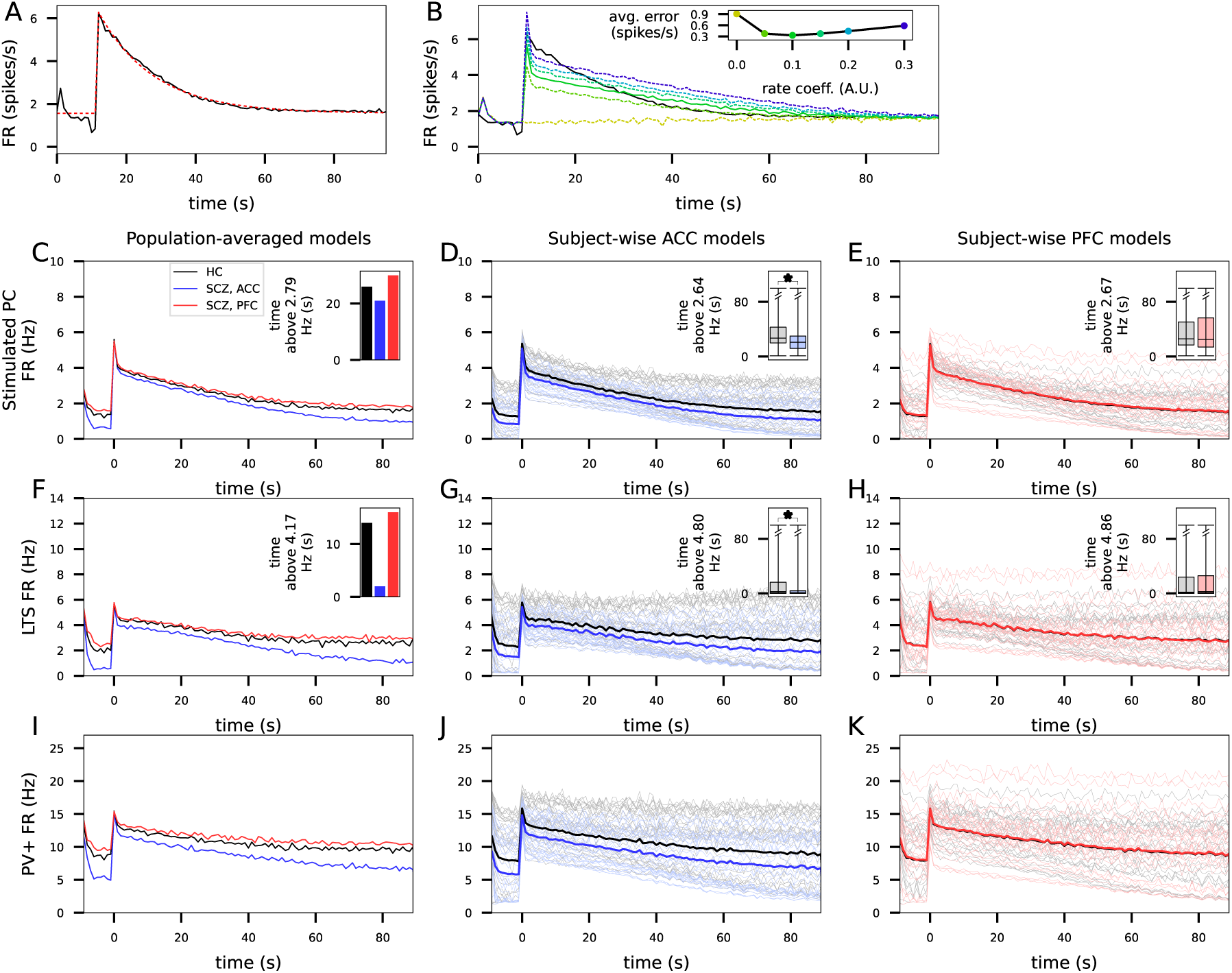
ACC-like variants of ion-channel expression in pyramidal cells and interneurons decrease the network response to external stimuli that mimic the increased firing caused by the WM-inducing stimulus. **A**: An exponential decay constant of 16 s fits the additional firing caused by the WM-induced stimulus in HCs. The black curve shows the FR of the stimulated pyramidal cells in the experiment of Fig. 3A, and the red dashed curve shows the best fit to the curve. **B**: The network with external WM-like stimulation with a decay constant of 16 s fit best to the default WM simulations when the additional input rate was 10% (at its peak) from the baseline input rate. The black curve shows the FR of the stimulated pyramidal cells in the experiment of Fig. 3A, and the coloured curves show the corresponding FR in the experiments where the HCN channel-modulating mGluR-mediated stimulus was replaced by an additional, external AMPAR-mediated synaptic stimulus that decayed with a decay constant *τ* =16 s and had a peak amplitude of 0% (no external stimulus; yellow dashed), 5% (green dashed), 10% (green), 15% (cyan dashed), 20% (light blue dashed), or 30% (blue dashed) of the baseline input rate. Inset: The average error in FR between default model and the model with external stimuli. The simulations with a 10% relative input rate fit best to the FR dynamics in the default network model of Fig. 3A. **C–M**: The simulations of Fig. 3 were repeated in the network where external WM-associated stimulus replaced the HCN channel-mediated persistent firing. See Fig. 3 for details.

**Figure S6:**
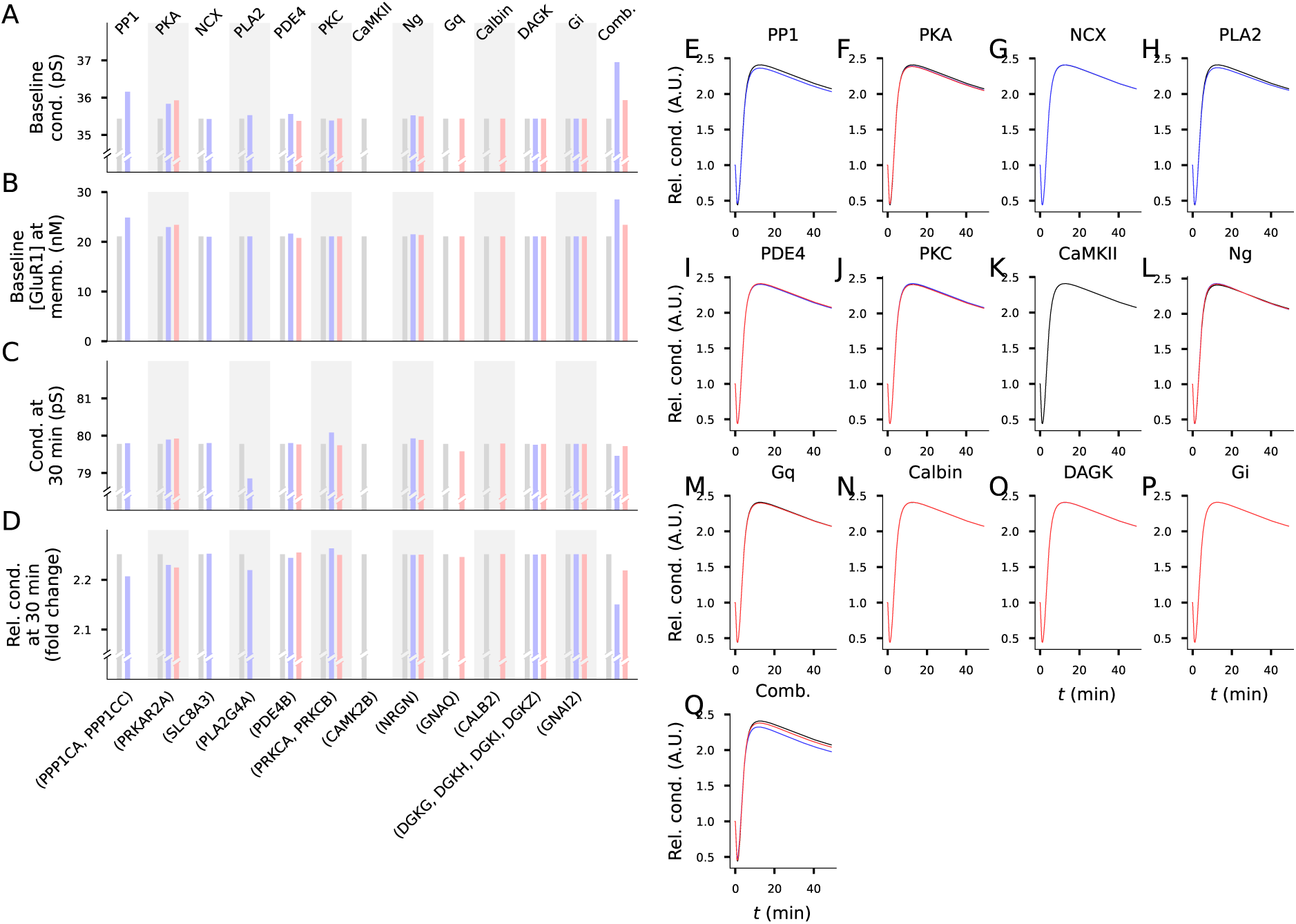
Altered expression of plasticity-regulating genes in SCZ affects the baseline synaptic conductance and decreases the amplitude of the potentiation at *t*=30 min after stimulus onset. A: The baseline synaptic conductance in the default model (representing HC) and in the models where expression of plasticity-regulating genes was altered according to SCZ data from ACC (blue) or PFC (red). The different columns represent different alterations of the model parameters as shown in Table 2: the concentration of PP1 was affected by expression levels of PPP1CA and PPP1CC in the ACC, PKA concentration by expression of PRKAR2A, NCX concentration by expression of SLC8A3 in the ACC, PLA2 concentration by expression of PLA2G4A, PDE4 concentration by expression of PDE4B, PKC concentration by expression of PRKCA in the ACC and PRKCB in the PFC, CaMKII concentration by expression of CAMK2B, Ng concentration by expression of NRGN, G-q concentration by expression of GNAQ, calbindin concentration by expression of CALB2 in the PFC, DAGK concentration by expression of DGKG and DGKI in the ACC, DGH in the PFC, and DGKZ in both ACC and PFC, and G-i concentration by the experession of GNAI2. The last column represents the data from the combined simulations where all expression level changes of Table 2 were implemented in the models. **B**: The baseline concentration of membrane-inserted GluR1 in the simulations of panel (A). **C**: The synaptic conductance 30 min after the onset of the 16 Hz*×*100 s stimulus in the simulations of (A). **D**: The relative (normalized by the baseline conductance shown in (A)) synaptic conductance 30 min after the stimulus onset. **E–Q**: The relative synaptic conductance time courses in the simulations of altered expression of plasticity-regulating genes.

**Figure S7:**
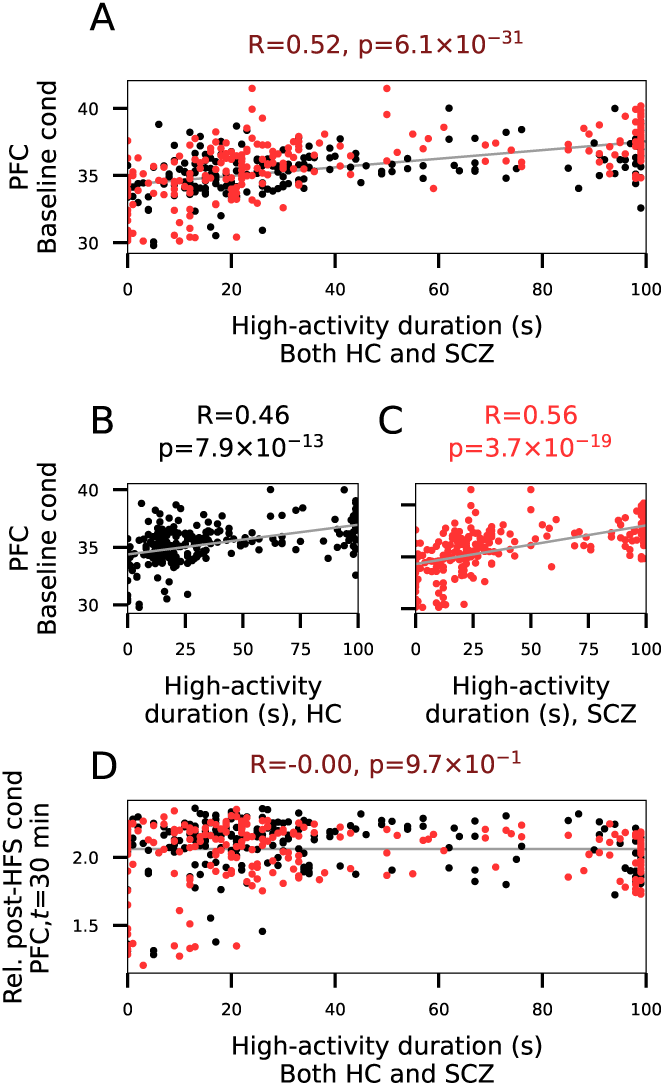
Correlations between alterations in the features of the predicted plasticity and the sustained firing in the PFC. **A–E**: The experiments of Fig. 5C–G repeated for the gene expression data from the PFC. See Fig. 5 for details.

**Figure S8:**
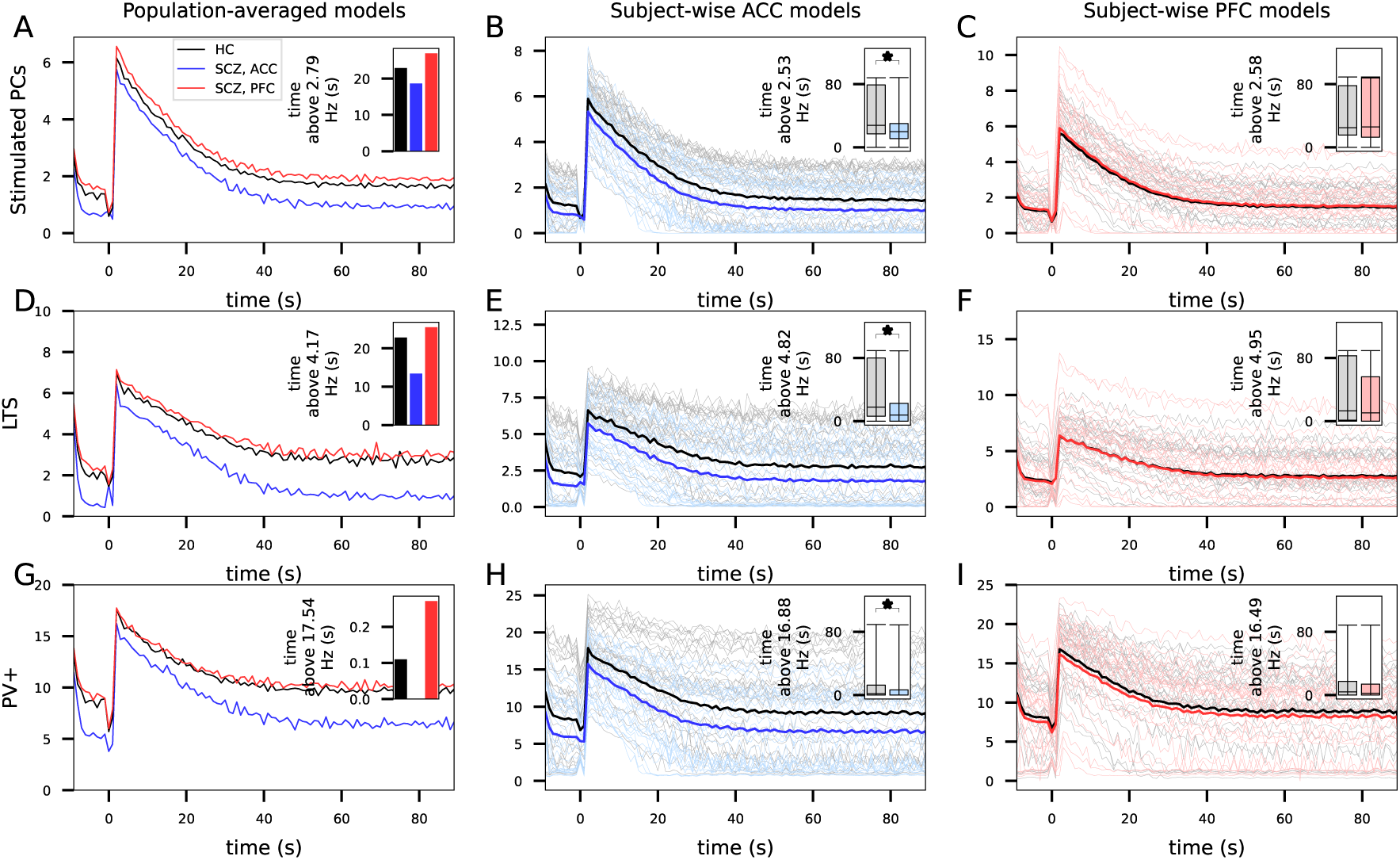
ACC-like variants of ion-channel expression in pyramidal cells and interneurons, combined with alterations of baseline synaptic conductance predicted by the biochemically detailed model [Mäki-Marttunen et al., 2024], decrease the network response to the WM-inducing stimulus. Here, all the simulations of Fig. S4 were repeated so that the baseline conductance of excitatory-to-excitatory synapses was altered according to the simulated data from Fig. 5. In the population-averaged simulations of (A), (D), and (G), the E*→*E synaptic conductance was 4.3% (ACC) or 1.4% (PFC) larger in SCZ than HC in addition to the parameter changes of Table 1, and in the personalized models (B–C), (E–F) and (H–I), the synaptic conductance was altered according to the subject-wise simulated data illustrated in the left insets of Fig. 5A–B. See Fig. S4 for details.

**Figure S9:**
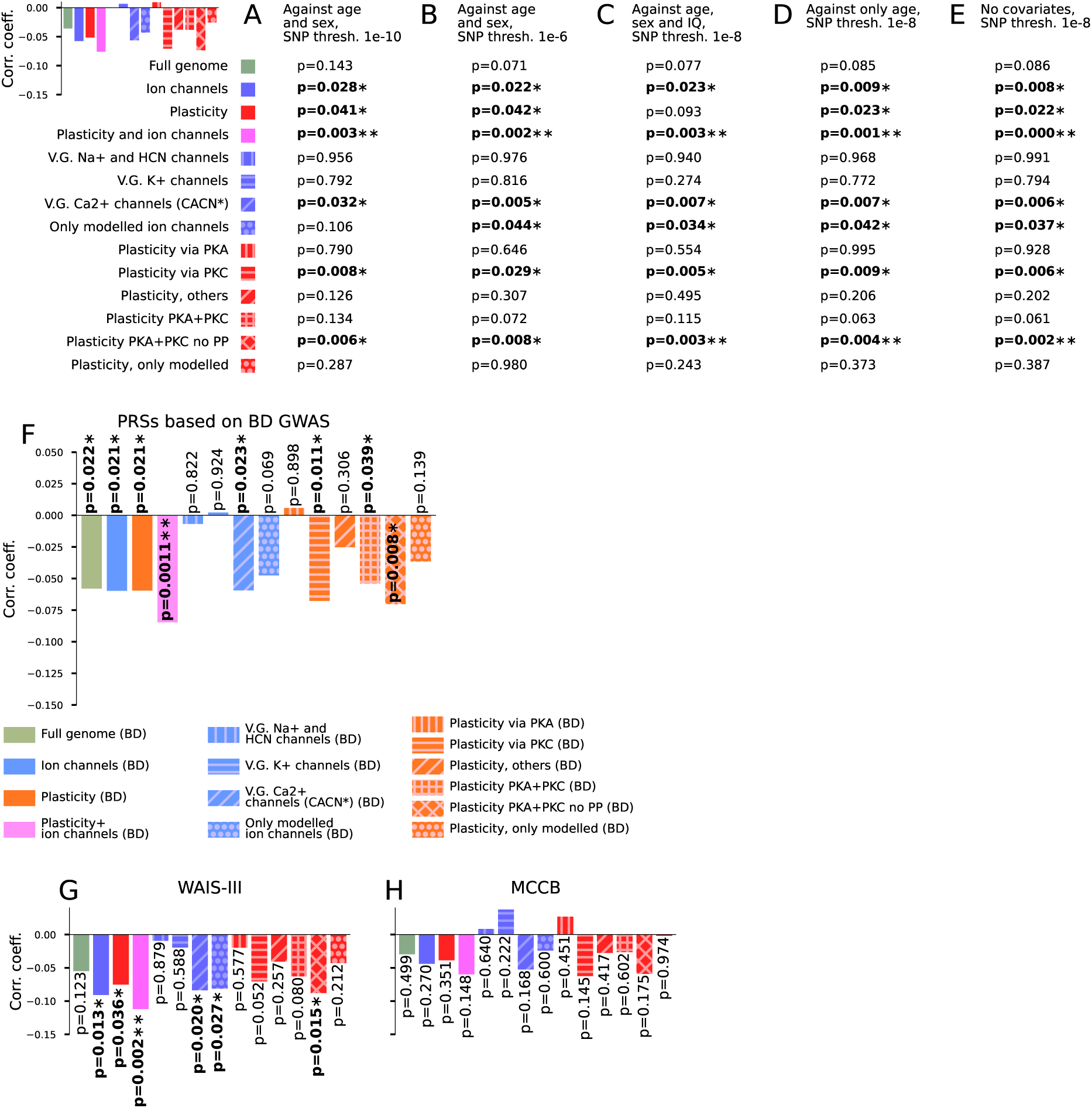
Results from the genotype-phenotype analysis are robust against differences in the set of covariates but depend on the cohort. Top left: The correlation coefficients of the PRSs and the WM performance (replotted from Fig. 6A). **A–B**: The analysis of Fig. 6A repeated using a p-value threshold of 1 *×* 10*^−^*^10^ (A) or 1 *×* 10*^−^*^6^ (B) for the SNP inclusion. See Fig. 6A for details. **C–E**: Results from the statistical tests when different sets of covariates were used. Statistically significant (p*<*0.05) associations marked with asterisks; the double asterisks indicate significance when Bonferroni-corrected for multiple tests (p*<*0.05/14). **F**: The analysis of Fig. 6A repeated using the BD-PRSs calculated from [O’Connell et al., 2025]. **G**: The analysis of Fig. 6A repeated using only the cohort where WAIS-III version of the LNS task was used. **H**: The analysis of Fig. 6A repeated using only the cohort where MCCB version of the LNS task was used.

**Figure S10:**
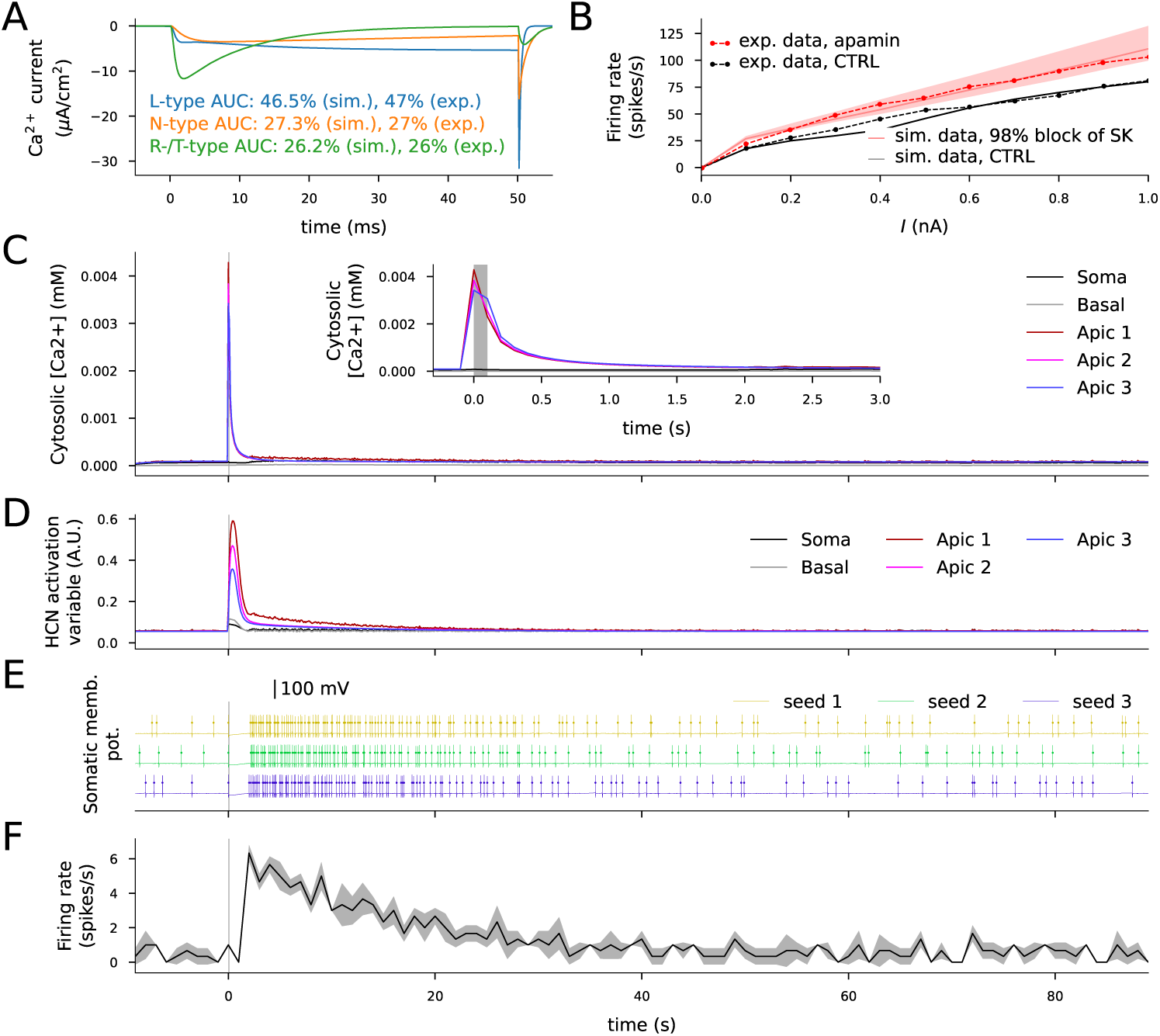
The pyramidal cells without a hot zone of Ca^2+^ channels behave similarly to those with the hot zone in response to somatic current injection but display a dampened intracellular Ca^2+^ response in the apical dendrite in response to the WM-inducing stimulus. A–F: The experiments of Fig. 8C–H repeated in pyramidal cells without the hot zone of Ca^2+^ channels. The voltage clamp response (A) and the f-I curves (B) were similar to those of the pyramidal cells with the hot zone of Ca^2+^ channels (Fig. 8C–D), but the WM-inducing stimulus caused a smaller Ca^2+^ response (C) and HCN channel activation (D) than in the pyramidal cells with the hot zone (Fig. 8E–F). This mildly increased the AP firing in response to the WM-like stimulus in the pyramidal cells without the hot zone of Ca^2+^ channels (E–F) compared to those with he hot zone (Fig. 8G–H).

